# Socioeconomic inequalities in smoking prevalence and intensity in Germany: A repeated cross-sectional analysis from 1998 to 2024

**DOI:** 10.64898/2026.06.12.26355514

**Authors:** Karl Emmert-Fees, Gesa Meyer, Michael Laxy, Michael Hanselmann

## Abstract

**Background:** Smoking inequalities by socioeconomic position (SEP) have widened consistently in Germany, but sex-specific trends beyond 2013 and inequalities in daily cigarette consumption among smokers (intensity) are unknown. We analyzed trends in absolute and relative socioeconomic inequalities in smoking prevalence and intensity among German adults across three decades.

**Methods:** We used 14 waves (1998-2024) of population-representative cross-sectional data from the German Socio-Economic Panel to estimate sex-specific trends in smoking prevalence and intensity in adults aged 25-64. Inequalities were quantified across strata of education, occupation, and equivalized household income using the absolute and relative concentration index with 95% bootstrap confidence intervals.

**Results:** Smoking prevalence declined from 35.05% (CI: 33.90%, 36.20% in 1998 to 22.19% (CI: 21.15%, 23.24%) in 2024, and mean intensity from 18 (CI: 17,18) to 13 (CI: 13, 14) cigarettes/day. Over this period sex-differences in both outcomes narrowed almost completely. Absolute and relative inequalities in smoking prevalence widened particularly for education and occupation. By 2024, the inequality gap between men and women has largely narrowed driven by a stagnating or rising smoking prevalence among low-SEP women (at least until 2018) alongside continued declines in higher-SEP women and for men. In fact, relative educational inequalities are now larger among women. Inequalities in smoking intensity, particularly related to income, were generally smaller than those in prevalence.

**Conclusion:** Socioeconomic smoking inequalities in Germany widened from 1998 to 2024 primarily driven by reductions among higher-SEP groups and increases in low-SEP women. Health equity must be considered in the German tobacco control strategy.

**Key messages:** *What is already known on this topic:*

– Socioeconomic inequalities in smoking, particularly according to education, have been increasing in Germany since the 1990s.
– Among men this increase was driven by reductions in smoking for those with a higher socioeconomic position, while among women, smoking prevalence has additionally increased for those with a lower socioeconomic position.
– It is unknown how inequalities in smoking prevalence and intensity according to education, occupation, and income have developed in Germany over the last 10 to 15 years.

*What this study adds:*

– Socioeconomic inequalities in smoking prevalence in Germany have continuously widened over almost three decades for both men (relative inequalities) and women (absolute and relative inequalities).
– Although sex differences in smoking prevalence inequalities have narrowed and have in some cases become larger among women compared to men, very recent trends may indicate a declining prevalence among women with a lower socioeconomic position.
– Socioeconomic inequalities in smoking intensity have also increased over time and are most relevant across strata of education and occupation.

*How this study might affect research, practice or policy:*

– Smoking is an important driver of health-related inequalities in Germany for women and men.
– Socioeconomic inequalities in smoking are substantial and there is a need for tobacco regulation that effectively reduces smoking overall and particularly among disadvantaged groups.
– Germany may be about to enter a new phase of the smoking epidemic where the prevalence of smoking among the most disadvantaged groups is starting to decline.

## INTRODUCTION

Tobacco smoking is an important risk factor responsible for a high preventable morbidity and mortality burden [1]. The detrimental health effects of smoking have been extensively documented [2,3]. Smoking increases the risk for many conditions, including cardiovascular diseases, respiratory diseases, and cancer, particularly of the lung [3]. It is estimated that globally, around 1.1 billion people currently smoke and that approximately 200 million disability-adjusted life years (DALYs) are attributable to smoking [1]. The World Health Organization (WHO) considers reducing smoking rates through evidence-based policy approaches a central goal for improving global population health. This has also been reflected in the 2003 WHO Framework Convention on Tobacco Control (FCTC) [4], which Germany signed in 2004.

Like other behavioral health risks, smoking follows a gradient along socioeconomic position (SEP), with a higher prevalence in more disadvantaged groups [5–8]. Beyond this gradient, dynamic smoking prevalence patterns by sex, age, and birth cohort have been observed over the past decades [6,8–14]. Socioeconomic inequalities in smoking over time are often conceptualized with the smoking epidemic model combined with sociological theories about the diffusion of innovations (i.e., smoking as a social practice) and the structural drivers of socioeconomic health inequalities [15–17]. For example, smoking was first common among affluent men before it became widespread among the working class. In the first half of the 19^th^ century, male smoking prevalence was as high as 80% in the US [11]. Patterns in other Western countries were likely similar despite a lack of historic data [10,12–14]. Women across socioeconomic strata picked up smoking 20-30 years later, partially linked to industry efforts to capitalize on first-wave feminism [18]. Trends are further shaped by initiation and quitting patterns. Smoking rates are typically the highest among young adults and decline with age [10,12,19]. While most people try cigarettes at least once in their lifetime, those from a disadvantaged background have fewer quitting attempts and quit less often successfully [20–23].

Germany is likely in a later stage of the smoking epidemic [12,24,25]. The overall prevalence of smoking has been declining for several decades but remains high compared to other high-income countries in Western Europe [12,26]. This is sometimes attributed to missing tobacco control policies [27]. According to estimates from 2022/2023, 33% of men and 25% of women currently smoke regularly [28]. Previous analyses documented a higher prevalence among those with a lower SEP according to education, occupation, and income [23–25,28–30]. Despite declining prevalence socioeconomic smoking inequalities among adults in Germany have widened over time, particularly for women [25,31,32]. Consistent with the smoking epidemic model, the observed increase in inequalities across education, occupation, and income have been driven by a largely stagnating prevalence among low-SEP men compared to decreases in high-SEP men [25]. Although smoking is also in decline among high-SEP women, prevalence among their low-SEP counterparts has been increasing [25].

However, recent sex-specific developments of smoking inequalities in Germany are unknown as analyses of long-term trends stop around 2014, use inconsistent sampling procedures over time, and lack a comprehensive assessment of different SEP dimensions [12,25,28]. Previous studies also did not analyze the number of tobacco products smoked among smokers (i.e., smoking intensity) which is important to fully capture the health burden of smoking [2,12,25,28].

To address these gaps, the objectives of this study are 1) to describe long-term trends and socioeconomic patterns in smoking prevalence according to education, occupation, and income by sex from 1998 to 2024; 2) to analyze whether trends and inequalities of smoking intensity have developed similarly to those of prevalence; and 3) to analyze whether the trend of increasing absolute and relative socioeconomic inequalities in smoking prevalence has continued into the 2020s.

## METHODS

### Study design & data

We conducted a repeated cross-sectional analysis of socioeconomic inequalities in smoking behavior, operationalized through smoking prevalence and intensity (i.e., number of cigarettes smoked per day among smokers), in German adults stratified by sex from 1998 to 2024. SEP was assessed according to education, occupation, and income which are dimensions commonly used to assess health inequalities [33]. We adhered to best practice guidance on conducting and reporting on observational studies (see *Data availability statement* and *Appendix 1*) [34].

We used repeated cross-sectional data from the German Socio-Economic Panel Survey (SOEP), an ongoing representative survey sampling private households in Germany. The widely used survey collects many socioeconomic and behavioral variables at the household and individual level [35]. In each yearly SOEP wave all adults (age 17+ years) of a sampled household answer individual questionnaires which in 1998, 1999, 2001 and every second year since 2002 also included instruments on smoking. However, the 1999 survey did not collect information on smoking intensity. To achieve consistency, we thus excluded the 1999 wave from this study and used data from the 14 waves that collected full smoking data between 1998 and 2024.

We restricted our analyses to adults aged 25 to 64, as younger adults may not have completed their education and may not yet be fully employed; older adults may already be retired and receive pension benefits. We further exclude individuals living in institutional settings.

### Smoking outcomes

To analyze smoking prevalence and intensity we used self-reported data. Since 2016 e-cigarettes and vapes are explicitly excluded in the wording of the respective questions (see details in *Appendix 2*). Respondents’ current smoking status was determined based on a binary question (e.g., “*Do you currently smoke, be it cigarettes, a pipe, or cigars?*”). Based on this, we constructed a consistent individual binary smoking status variable.

Smoking intensity was assessed through the self-reported daily consumption of tobacco products among respondents that reported to be current smokers (e.g., “*How many cigarettes, pipes, or cigars do you smoke per day?*”). We directly used the response scale (in *cigarettes/day* for simplicity) as our continuous smoking intensity outcome.

### Socioeconomic dimensions

Education was defined by educational attainment (i.e., highest educational degree achieved) using four levels aggregated from the Comparative Analysis of Social Mobility in Industrial Nations (CASMIN) classification [36]: *No graduation or basic* – individuals with no graduation or completed elementary education (CASMIN categories: 1a, 1b, 1c); *Intermediate* – individuals with intermediate general education (2a, 2b); *Higher* – individuals with general or vocational maturity (2c_gen, 2c_voc); and *Academic* – individuals with tertiary education (3a, 3b).

Occupation was defined by the last reached occupational status using the Socio-Economic Index of Occupational Status (ISEI) [37]. Thus, this does neither capture current employment status or length of unemployment. For participants that never practiced any profession, the ISEI is missing. We do not assess the unemployed as a separate occupational group. ISEI scores range from 16 (lowest, e.g. support staff or cleaning personnel) to 90 (highest, e.g. judges). Based on the yearly ISEI distribution, we divided our sample into five quintiles, which roughly correspond to common occupational (see *Appendix 2*).

Income was defined by the annual equivalized net household income which we computed based on total household net income and household structure following standard OECD methodology [38]. The household head received a weight of 1.0, each additional adult a weight of 0.5, and each child a weight of 0.3. Using the annual distribution of equivalized household income, we divided the sample into five income quintiles.

### Measuring health inequalities

To analyze socioeconomic inequalities in smoking behavior over our study period, we calculated the Absolute (ACI) and Relative Concentration Index (RCI) for each year and socioeconomic dimension [39,40]. The ACI is a standardized absolute measure of inequality based on the covariance between the cumulative population share of an inequality measure and an outcome [39,40]. Advantages and disadvantages of the ACI compared to other measures are discussed in *Appendix 2*.

The ACI operates on the scale of the respective outcome. The RCI is the relative counterpart and ranges from −1 to +1. It scales the ACI by the mean of the outcome. Positive values indicate that the health outcome is disproportionately concentrated among those with higher SEP, whereas negative values indicate the opposite [39,40]. Thus, for both smoking outcomes in our study decreasing (i.e., more negative) values over time would indicate increasing inequalities.

### Statistical analysis

We estimated smoking prevalence and intensity stratified by sex and each socioeconomic dimension following a descriptive rationale. Smoking prevalence was calculated as the annual proportion of current smokers. Smoking intensity was calculated as the mean daily number of tobacco products smoked among smokers each year.

We assessed statistical uncertainty through 95%-confidence intervals (CI) based on empirical standard errors. All estimations were conducted using the individual annual cross-sectional weights provided in the SOEP dataset, which adjust for unit non-response and deviations from the population structure [35,41,42]. As a robustness check, we additionally estimated age-standardized smoking prevalence using the 2024 population as a reference (*Appendix 3*). We explicitly opted against using age-standardization as the main analysis to analyze actual trends over time. We additionally conducted exploratory analyses to examine interactions between education, occupation, and income (*Appendix 4*). Based on the yearly smoking prevalence and intensity values, we calculated the ACI and RCI for each socioeconomic dimension stratified by sex. Here, uncertainty was estimated with a bootstrap approach with 1,000 replications (*Appendix 2*).

All analyses were performed using an analysis-wise complete case approach. Thus, sample sizes vary across analyses (*Figure A1*). We opted for this approach because response rates in the SOEP are generally high. In fact, for selected variables (e.g., income) officially imputed versions are provided [35,43]. The proportion of missing data for all variables included in the analysis is reported in *Table A1*.

All analyses were performed with *R* (version 4.5.0) and particularly the *survey* package [44]. All analysis code is publicly available at https://osf.io/ex75z/.

## RESULTS

### Sample characteristics

Our analytical sample comprised 222,898 observations. The proportion of missing data per variable was very low except for the ISEI and education, for which approximately 16.14% respectively 5.27% of data was missing (*Table A1*). *Table 1* presents socio-demographic characteristics in 1998 and 2024.

**Table 1:** Sample demographics for the SOEP waves of 1998 and 2024.

|  | 1998<br>(n* = 10521) |  | 2024<br>(n* = 20192) |  |
| --- | --- | --- | --- | --- |
|  | Estimate | (Std. Dev.) | Estimate | (Std. Dev.) |
| **Sex** ( = female) | 49.45% |  | 49.39% |  |
| **Age** in years (Mean) | 44.15 | (0.14) | 45.45 | (0.15) |
| **Migration Background** (= yes) | 16.54% |  | 32.67% |  |
| **Relationship Status** |  |  |  |  |
| Married or Registered Partnership | 70.41% |  | 56.99% |  |
| Widowed | 3.04% |  | 1.57% |  |
| Divorced or Separated | 9% |  | 10.46% |  |
| Single, Never Married | 17.55% |  | 30.99% |  |
| **Educational Level** |  |  |  |  |
| No graduation or Basic | 48.34% |  | 19.5% |  |
| Intermediate | 25.55% |  | 24.17% |  |
| Higher | 8.62% |  | 19.25% |  |
| Academic | 17.49% |  | 37.08% |  |
| **Occupational Status (ISEI)** |  |  |  |  |
| Overall (Mean ISEI Score) | 44.15 | (0.21) | 52.58 | (0.28) |
| 1. Quintile - lowest | 25.34 | (0.12) | 22.38 | (0.15) |
| 2. Quintile - low | 34.9 | (0.07) | 39.95 | (0.21) |
| 3. Quintile - medium | 43.13 | (0.11) | 55.32 | (0.07) |
| 4. Quintile - high | 53.21 | (0.06) | 68.23 | (0.14) |
| 5. Quintile - highest | 68.81 | (0.21) | 79.91 | (0.11) |
| **Income\*\*** |  |  |  |  |
| Overall (Mean) | 17260.6 | (117.41) | 34738.73 | (280.68) |
| 1. Quintile - lowest | 8520.13 | (59.39) | 13505.48 | (144.79) |
| 2. Quintile - low | 12623.64 | (21.69) | 24106.12 | (57.19) |
| 3. Quintile - medium | 15527.45 | (25.27) | 30940.71 | (54.41) |
| 4. Quintile - high | 19530.18 | (39.64) | 38812.27 | (73.52) |
| 5. Quintile - highest | 30113.79 | (335.16) | 66339.45 | (919.29) |

**Notes:** The table presents sociodemographic characteristics for the SOEP waves of 1998 and 2024. Cross-sectional weights were applied to ensure representativeness for the German population between the age of 25 to 64. \* The presented n is the size of the respective analytical SOEP wave sample. Due to missing data for some variables, the numbers that are presented in the table are based on differing sample sizes. To get an impression about the share of missing data for each variable please refer to Table A1 in Appendix 3. \*\*Annual equivalized net household income.

### Smoking prevalence and intensity

We find that from 1998 to 2024 smoking prevalence in Germany among adults aged 25 to 64 has continuously declined from 35.05% (CI: 33.90%, 36.20%) to 22.19% (CI: 21.15%, 23.24%) (*Table 2*, *Figure A2)*. While in 1998 smoking prevalence was around 10%-points lower among women than men, this difference has largely disappeared in 2024 and continuously narrowed over time (*Table 2*, *Figure A2)*.

**Table 2:**
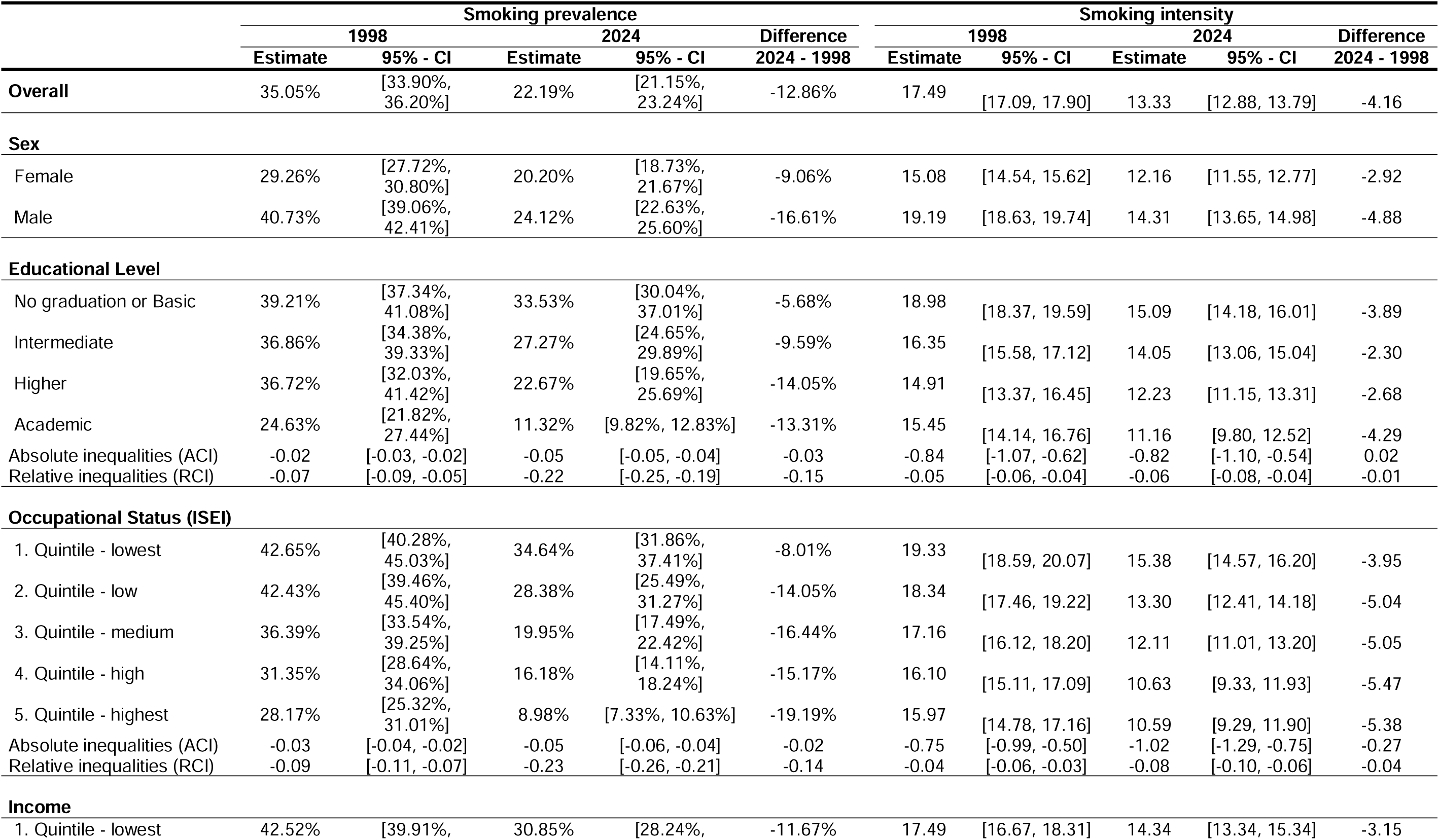
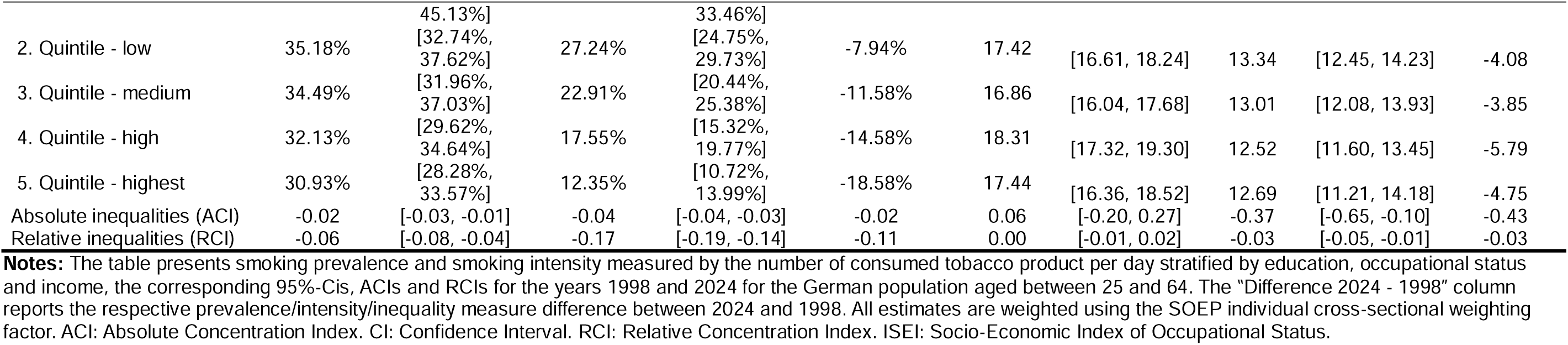
Socioeconomic inequalities in smoking prevalence and intensity for 1998 and 2024.

Over the same period smoking intensity has also declined from 17.49 cigarettes/day (CI: 17.09, 17.90) to 13.33 cigarettes/day (CI: 12.88, 13.79) (*Table 2*, *Figure A3*). Men on average smoked 4 cigarettes more per day than women in 1998. Until 2024 this gap has continued to narrow substantially (*Table 2*, *Figure A3*).

### Educational inequalities

Population-wide absolute and relative educational inequalities in smoking prevalence have increased substantially between 1998 and 2024 and these trends are particularly driven by women (*Figure 1*, *Tables A3*). In fact, among women with basic education the share of smokers has increased by about 4%-points between 1998 and 2024 whereas it has declined by about 15%-points for men. In contrast, the prevalence has roughly halved among men and women with academic degrees (*Figure 1*, *Table A3, Table A4*). Both absolute and relative educational inequalities have increased for women from 1998 (ACI –0.01, CI: –0.02, –0.00; RCI –0.03, CI: – 0.06, –0.00) to 2024 (ACI –0.05, CI: –0.06, –0.04; RCI –0.24, CI: –0.29, –0.20) (*Figure 3*, *Table A3*). Among men, absolute educational inequalities changed little, while relative inequalities increased markedly (RCI –0.09 [CI: –0.12, –0.07] to –0.20 [CI: –0.24, – 0.16]) (*Table A4*). Thus absolute inequalities have converged and relative inequalities are now larger for women (*Figure 3*) even though smoking among women with lower education has started to decline since 2018 (*Figure 1*).

**Figure 1:**
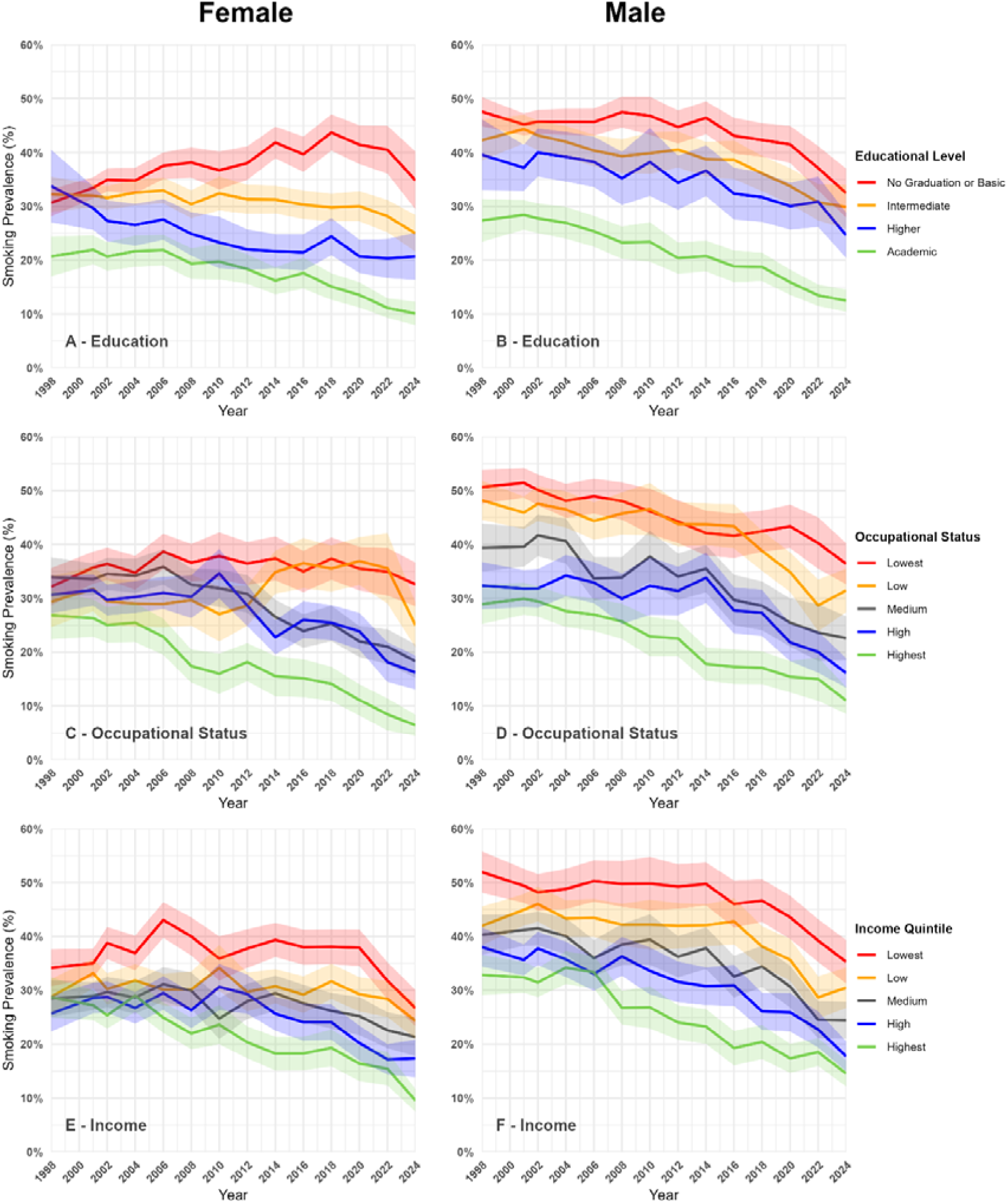
Smoking prevalence according to socioeconomic indicators from 1998 to 2024 for men and women. **Notes:** Data: SOEP v41. Sample: adults aged 25-64. Lines show estimated smoking prevalence by education, occupational status and income, separately for women and men, 1998-2024. Education levels follow the CASMIN classification (please see the methods section for details); occupational status (ISEI) and equivalized net household income (Income) are split into population-weighted quintiles for each survey year. Shaded bands represent 95% confidence intervals. All estimates are weighted using the SOEP individual cross-sectional weighting factor.

Educational inequalities in smoking intensity are smaller than those related to prevalence and in 2024 smokers with basic education smoked around 15 cigarettes/day (CI: 14, 16) compared to 11 cigarettes/day (CI: 10, 13) for the highly educated (*Table 2*, *Figure A3*). Educational inequalities in smoking intensity were historically higher among men, but sex differences in absolute and relative inequalities have now narrowed (*Figure 2*, *Table A3, Table A4, Figure A4*).

**Figure 2:**
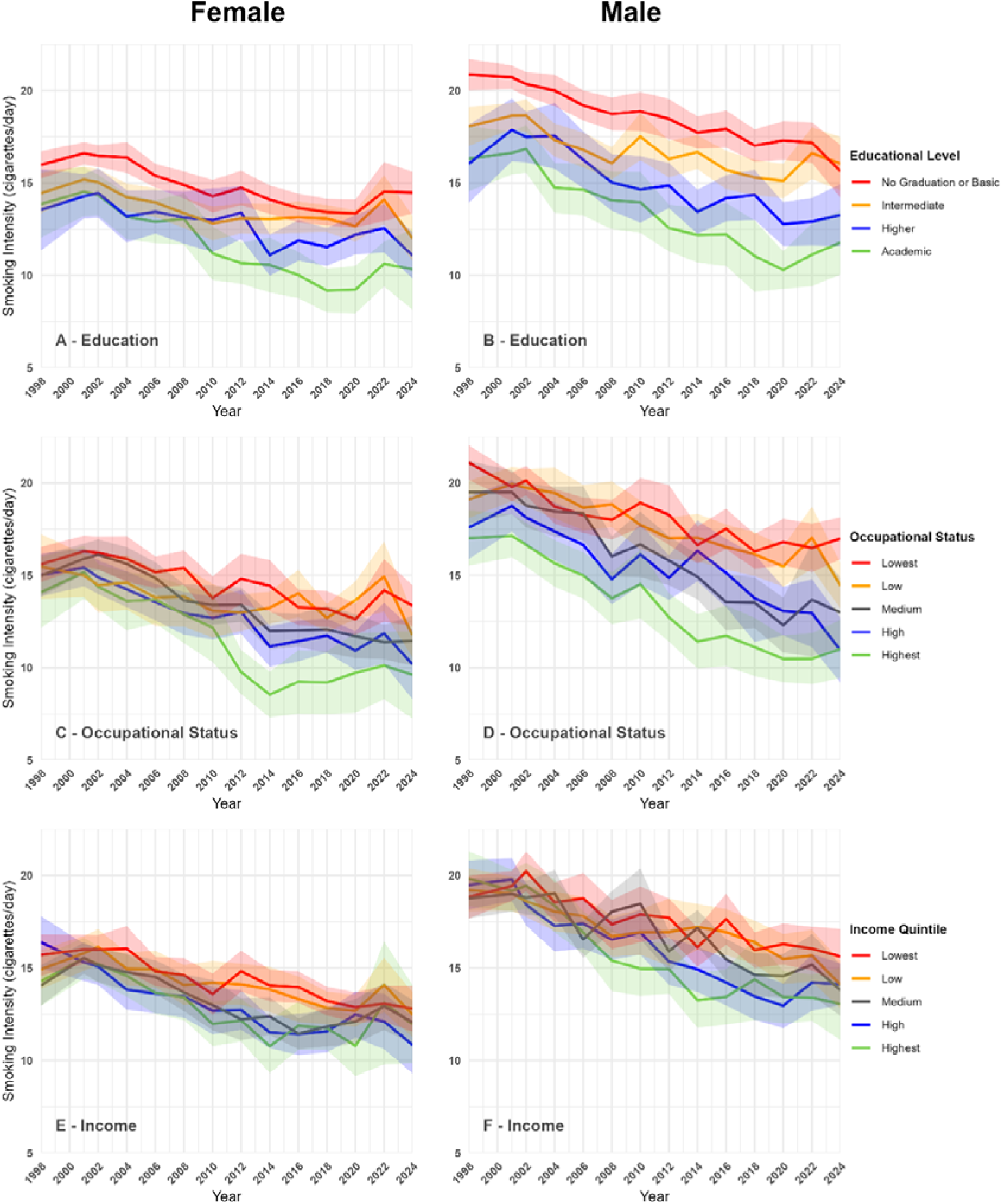
Smoking intensity according to socioeconomic indicators from 1998 to 2024 for men and women. **Notes:** Data: SOEP v41. Sample: current smokers aged 25-64. Lines show mean self-reported daily cigarette consumption (smoking intensity) by education, occupational status and income, separately for women and men, 1998-2024. Education levels follow the CASMIN classification (please see the methods section for details); occupational status (ISEI) and equivalized net household income (Income) are split into population-weighted quintiles for each survey year. Shaded bands represent 95% confidence intervals. All estimates are weighted using the SOEP individual cross-sectional weighting factor.

### Occupational Inequalities

Population-wide absolute and relative occupational inequalities in smoking have also widened considerably between 1998 and 2024 despite differential underlying sex trends (*Figure 1*, *Table 2*, *Figure A2)*. Among women, smoking prevalence remained roughly stable in the lowest occupational quintile but declined sharply in higher-status quintiles, while among men we observed steady decreases across the whole distribution (*Figure 1*, *Table A3, Table A4)*. As a result, although inequality levels have increased for women and stayed approximately constant for men between 1998 and 2024 (*Figure 3*), the gradient between women (ACI –0.05, CI: –0.06, –0.04; RCI –0.23, CI: –0.27, –0.20) and men (ACI –0.05, CI: –0.06, –0.05; RCI –0.23, CI: –0.26, –0.19) has narrowed substantially (*Figure 3*, *Table A3*, *Table A4*).

Occupational inequalities in smoking intensity are larger among men compared to women but roughly doubled for the latter over the study period (*Figure 2*, *Table A3*, *Figure A4)*. In 2024 smokers in the lowest ISEI quintile smoked 15 cigarettes/day (CI: 15, 16) compared to 11 cigarettes/day (CI: 9, 12) for those in the highest quintile (*Table 2*, *Figure A3)*.

### Income-related inequalities

Income-related inequalities in smoking have also increased between 1998 and 2024 but are slightly lower than those found for education and occupation albeit with comparable sex patterns (*Figure 1*, *Table 2*, *Figure A2)*. Similar to education and occupation, the sex gradient in income-related smoking prevalence inequalities has narrowed leading to persistent inequalities for men (ACI –0.04, CI: –0.05, –0.04; RCI – 0.18, CI: –0.21, –0.15) and women (ACI –0.03, CI: –0.04, –0.02; RCI –0.16, CI: –0.19, – 0.12) in 2024 (*Figure 3*, *Table A3, Table A4)*.

**Figure 3:**
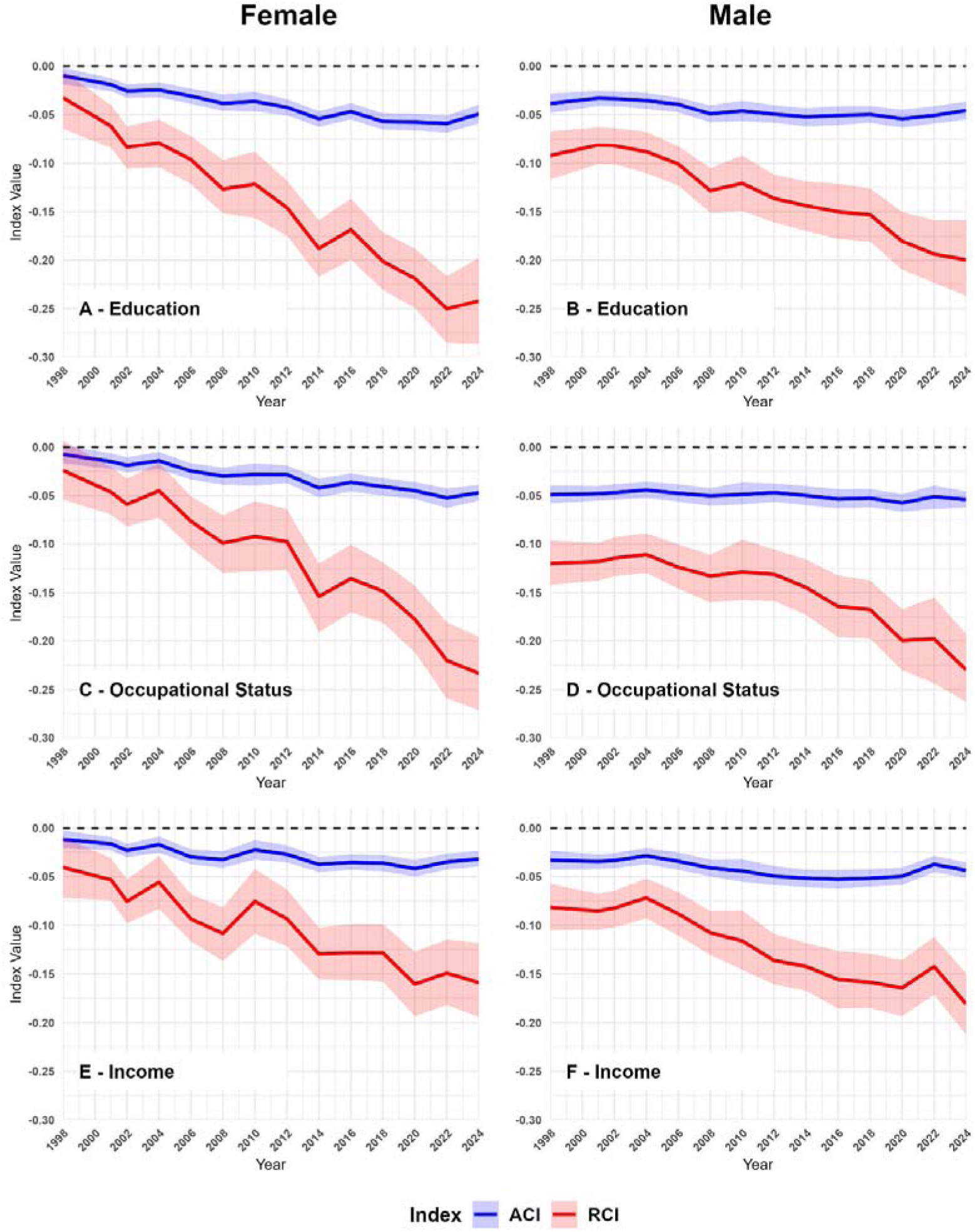
Absolute and relative inequalities in smoking prevalence according to socioeconomic indicators from 1998 to 2024 for men and women. **Notes:** Data: SOEP v41. Sample: adults aged 25-64. Lines show the absolute (ACI) and relative (RCI) concentration index for smoking prevalence by education, occupational status and income over time, separately for women and men. Negative values indicate higher smoking prevalence among individuals with lower socio-economic status. Education levels follow the CASMIN classification (please see the methods section for details); occupational status (ISEI) and equivalized net household income (Income) are split into population-weighted quintiles for each survey year. Confidence intervals (shaded bands) are 95% percentile intervals from 1,000 bootstrap replications. All estimates are weighted using the SOEP individual cross-sectional weighting factor.

We observed a sharp drop in smoking prevalence among households in the lower income quintiles after 2020. This could be the result of actual reductions in smoking or shifts in the income distribution as a result of labor market shocks during the COVID-19 pandemic [45] which increased the proportion of highly educated in lower income quintiles (*Figures A11 – A14*). Income-related inequalities in intensity are small increasing somewhat since 1998 for women and increasing substantially for men (*Figure 2*, *Table 2*, *Figure A3, Figure A4)*.

### Robustness checks and exploratory analyses

We observe barely any difference between standardized and non-standardized analyses, except for some educational groups in 1998 (*Table A2*). Exploratory analyses show that particularly educational gradients persist across strata of the other SEP dimensions (*Figures A5-A10*).

## DISCUSSION

Using representative survey data spanning 1998 to 2024, we analyzed long-term trends in socioeconomic inequalities in smoking prevalence and intensity among German adults aged 25-64. We find that smoking prevalence declined substantially over the study period and that smoking intensity among smokers likewise decreased, while the sex gap in both outcomes narrowed substantially. However, population-level absolute and relative inequalities in smoking prevalence consistently widened, with the steepest increase observed for education and occupational status. These developments are primarily driven by stagnating or increasing smoking prevalence among low-SEP women. Since 1998 sex differences in socioeconomic inequalities have also converged across all three dimensions. Relative educational inequalities are now larger for women. Yet, the most recent data shows that smoking prevalence among low-SEP women has begun to decline substantially, a pattern consistent with the later stages of the smoking epidemic model [15,16]. Socioeconomic inequalities in smoking intensity followed largely parallel but generally more modest trends.

Our findings are broadly consistent with previous national and international studies on long-term trends in socioeconomic smoking inequalities [12,24,25,28,30,46]. Using Microcensus data from 1995 to 2013, Tönnies et al. found that both absolute and relative inequalities in smoking prevalence increased across education, income, and occupation groups with larger increases observed among women than men [25]. Kuntz et al. found widening occupational differences in smoking among employed adults from 1999 to 2013 [30]. Heilert and Kaul came to similar conclusions analyzing SOEP data from 1998 to 2014 [12]. We largely confirm these patterns and demonstrate that (relative) socioeconomic inequalities have widened particularly for women. A narrowing of the sex gap in inequalities was already emerging in previous studies [25] but partly contrasts with Hoebel et al. [24], who reported a statistically significant increase in relative educational inequalities only for men from 2003 to 2012. These discrepancies are plausibly related to different comparison timepoints [24]. Importantly, our analysis suggests that smoking prevalence among less educated women may have indeed begun to decline from ∼43% in 2018 to ∼35% in 2024 in this group. This could be indicative of a transition to later stages of the smoking epidemic in Germany but also plausibly related to changes in demographic composition due to migration [15,16]. Our results partially contradict data reported in a recent analysis from Starker et al. (2025) that shows only small increases in absolute educational differences in smoking since the early 2000s and does not evaluate sex-specific trends [28]. Our results are consistent with analyses from other high-income countries which show that declines in overall smoking prevalence have been accompanied by stable or widening (relative) socioeconomic inequalities, with the residual burden increasingly concentrated among the most disadvantaged. Long-term studies converge on this pattern, regardless of the SEP dimension or inequality metric used [5,8,9,14,47–49].

In contrast to prevalence smoking intensity has rarely been studied previously in Germany and never with formal inequality metrics. Heilert and Kaul reported a descriptive SEP gradient in cigarettes smoked per day [12]. We show that intensity inequalities have consistently widened over time and that there are unexpectedly small income-related intensity inequalities which is striking given that costs of smoking directly scale with intensity and has implications for tobacco regulation in Germany. The few available international analyses that report inequalities in smoking intensity (e.g., from China [7]) likewise show a clear SEP gradient.

Our analysis has several important strengths. We covered almost three decades of data using a high-quality nationally representative survey and conducted a comprehensive assessment of socioeconomic inequalities in smoking behavior in Germany by assessing both smoking prevalence and intensity. We assessed inequalities according to three important SES dimensions and used the ACI and RCI as established metrics. These indices are also particularly useful for trend comparisons. Our rigorous approach visualizes long-term trends and enables a detailed assessment of the contemporary phase of the smoking epidemic in Germany.

Important limitations of our study include that we use self-reported data which is prone to reporting biases including social desirability bias. As long as these do not change over time, long-term comparisons remain valid. However, the socio-cultural relevance of smoking has changed since the late 1990s [15,18]. Yet we consider it to be implausible that such effects would fully explain the qualitative conclusions we draw. Additionally, SOEP contains neither information on the age at which participants started or quit smoking, nor on quitting attempts. We did also not assess inequalities related to e-cigarettes and vapes because these are rarely consumed among SOEP participants (data not shown). Finally, unlike other metrics, the ACI does not reflect the predicted gap between extreme SEP groups and may not be intuitively interpretable. Furthermore, we observed increasing rates of missing data for education in 2022 (14.15%) and 2024 (24.13%). According to communication with the SOEP team this is a known data gap in recent refreshment samples.

Our study contributes to tracking long-term trends in smoking inequalities, which is important for health policy given the health-damaging effects of tobacco, its contribution to health inequalities, and government efforts to regulate tobacco through policies [2,4,27,50]. In 2021, Germany ranked 34 of 37 according to the Tobacco Control Scale (TCS), which tracks implementation of FCTC recommendations across European countries [27]. Although some measures – such as place-based bans, warning labels, and tax increases – have been implemented, overall tobacco control largely focus on individual responsibility for behavior change [12]. However, this focus implies approaches that rely on individual agency and thus personal cognitive, sociocultural, temporal, and financial resources to act on health information, which are unequally distributed along SEP dimensions. Indeed, the results of our analysis support the consequent concern that individuals with a higher SEP react faster and more strongly to high-agency tobacco control policies, which steepens socioeconomic smoking gradients [15,51]. This makes structural (low-agency) policies that change the societal conditions for health behavior the primary choice to reduce inequalities but produces a challenge for public health policy because these approaches are often conceived paternalistic and require cross-sector collaboration [52,53]. Given this, it may seem unintuitive that smoking prevalence was less driven by income (*Figures A5 & A6*). However, this is likely explained by low-income smokers shifting to roll-your-own tobacco, which is less affected by current taxation in Germany and much cheaper per cigarette [54–56]. Consistently applied prohibitive pricing could thus support declines in smoking among disadvantages groups which carry the primary health burden of smoking [14,26,27,55,56].

Future studies should aim to quantify smoking inequalities more granularly to identify socioeconomic sub-groups that have the highest smoking prevalence and would benefit the most from targeted policies. To better understand the societal burden of smoking in Germany it is also important to update and accurately quantify the morbidity, mortality, and healthcare costs related to smoking. Lastly, it is paramount to better understand the behavioral mechanisms and combinations of policies that are effective in reducing both smoking overall and related socioeconomic inequalities.

## CONCLUSION

Driven by female smoking, socioeconomic inequalities in smoking prevalence in Germany have widened from 1998 to 2024, despite substantial declines in overall prevalence. Inequalities are now similar in women and men across all three SES dimensions. Early signs of a decrease in smoking among less educated women after 2018 may signal a transition into a later stage of the smoking epidemic. At the same time, the persistent lack of an income gradient in smoking intensity points to limits of current price-based measures alone. Closing these gaps will require stronger and more equity-oriented tobacco control in Germany that complements taxation with other structural policy approaches recommended within the FCTC.

## Acknowledgements

An early version of this analysis was conducted as a Master’s Thesis by Gesa Meyer during her studies at the Technical University of Munich.

## Contributors

MH had the initial idea for the study. MH, KEF, GM, and ML conceived and designed the study. MH and GM conducted the data preparation. MH and KEF performed the statistical analyses. Anthropic Claude Opus 4.6 and OpenAI ChatGPT 5.4 were used to implement, debug, and improve parts of the analysis code. KEF drafted the manuscript. GM, MH, ML, and KEF critically revised the manuscript for important intellectual content. Anthropic Claude Opus 4.6 was used to critically revise sections of the manuscript with the goal of improving reporting clarity. All authors approved the final version of the manuscript. KEF is the guarantor and accepts full responsibility for the finished work and the conduct of the study, had access to the data, and controlled the decision to publish.

## Patient consent for publication

Not applicable.

## Ethics approval

This study is a secondary analysis of anonymized data from the German Socio-Economic Panel (SOEP) study. Ethical clearance and informed consent for data collection were obtained by the German Institute for Economic Research (DIW Berlin) at the time of data collection. Thus, no separate ethics approval was required for this analysis. Further information is available at https://www.diw.de/soep.

## Data availability statement

The data of the German Socio-Economic Panel (SOEP) study is available for academic research purposes upon request at https://www.diw.de/en/diw_01.c.601584.en/data_access.html. The complete analysis code is publicly available at https://osf.io/ex75z/.

