## Appendix for "Socioeconomic inequalities in smoking prevalence and intensity in Germany: A repeated cross-sectional analysis from 1998 to 2024"

**Socioeconomic inequalities in smoking behavior in Germany: A descriptive trend analysis from 1998 to 2024**

### Appendix 1: STROBE Checklist

|  | Item No. | Recommendation | Section | Paragraph |
| --- | --- | --- | --- | --- |
| **Title and abstract** | 1 | (*a*) Indicate the study’s design with a commonly used term in the title or the abstract | Title | n/a |
|  |  | (*b*) Provide in the abstract an informative and balanced summary of what was done and what was found | Abstract | n/a |
| Introduction | | | |  |
| Background/rationale | 2 | Explain the scientific background and rationale for the investigation being reported | Introduction | Paragraphs 1-4 |
| Objectives | 3 | State specific objectives, including any prespecified hypotheses | Introduction | Paragraph 5 |
| Methods | | | |  |
| Study design | 4 | Present key elements of study design early in the paper | Methods – Study design & data | Paragraph 1 |
| Setting | 5 | Describe the setting, locations, and relevant dates, including periods of recruitment, exposure, follow-up, and data collection | Methods – Study design & data | Paragraphs 1-2 |
| Participants | 6 | (*a*) *Cohort study*—Give the eligibility criteria, and the sources and methods of selection of participants. Describe methods of follow-up  *Case-control study*—Give the eligibility criteria, and the sources and methods of case ascertainment and control selection. Give the rationale for the choice of cases and controls  *Cross-sectional study*—Give the eligibility criteria, and the sources and methods of selection of participants | Methods – Study design & data | Paragraphs 2-3 |
|  |  | (*b*) *Cohort study*—For matched studies, give matching criteria and number of exposed and unexposed  *Case-control study*—For matched studies, give matching criteria and the number of controls per case | n/a | n/a |
| Variables | 7 | Clearly define all outcomes, exposures, predictors, potential confounders, and effect modifiers. Give diagnostic criteria, if applicable | Methods – Smoking outcomes; Methods – Socioeconomic dimensions | All paragraphs |
| Data sources/ measurement | 8* | For each variable of interest, give sources of data and details of methods of assessment (measurement). Describe comparability of assessment methods if there is more than one group | Methods – Smoking outcomes; Methods – Socioeconomic dimensions; Appendix 2 | All paragraphs |
| Bias | 9 | Describe any efforts to address potential sources of bias | Methods – Statistical analysis | Paragraph 2-3 |
| Study size | 10 | Explain how the study size was arrived at | Methods, Appendix 4 | n/a |
| Quantitative variables | 11 | Explain how quantitative variables were handled in the analyses. If applicable, describe which groupings were chosen and why | Methods – Smoking outcomes; Methods – Socioeconomic dimensions | All paragraphs |
| Statistical methods | 12 | (*a*) Describe all statistical methods, including those used to control for confounding | Methods – Statistical analysis; Appendix 2 | Paragraphs 1-2 |
|  |  | (*b*) Describe any methods used to examine subgroups and interactions | Methods – Statistical analysis | Paragraphs 1-2 |
|  |  | (*c*) Explain how missing data were addressed | Methods – Statistical analysis | Paragraph 3 |
|  |  | (*d*) *Cohort study*—If applicable, explain how loss to follow-up was addressed  *Case-control study*—If applicable, explain how matching of cases and controls was addressed  *Cross-sectional study*—If applicable, describe analytical methods taking account of sampling strategy | Methods – Statistical analysis | Paragraph 2 |
|  |  | (*e*) Describe any sensitivity analyses | Methods – Statistical analysis | Paragraph 2 |
| **Results** | | |  |  |
| Participants | 13 | (a) Report numbers of individuals at each stage of study—eg numbers potentially eligible, examined for eligibility, confirmed eligible, included in the study, completing follow-up, and analysed | Results – Sample characteristics; Appendix 4 | All paragraphs |
|  |  | (b) Give reasons for non-participation at each stage | Results – Sample characteristics; Appendix 4 | All paragraphs |
|  |  | (c) Consider use of a flow diagram | Results – Sample characteristics; Appendix 4 | All paragraphs |
| Descriptive data | 14 | (a) Give characteristics of study participants (eg demographic, clinical, social) and information on exposures and potential confounders | Results – Sample characteristics; Table 1; Appendix 3 | All paragraphs |
|  |  | (b) Indicate number of participants with missing data for each variable of interest | Results – Sample characteristics; Appendix 3 | All paragraphs |
|  |  | (c) *Cohort study*—Summarise follow-up time (eg, average and total amount) | n/a | n/a |
| Outcome data | 15 | *Cohort study*—Report numbers of outcome events or summary measures over time  *Case-control study—*Report numbers in each exposure category, or summary measures of exposure  *Cross-sectional study—*Report numbers of outcome events or summary measures | Results – Smoking prevalence and intensity; Table 2 | All paragraphs |
| Main results | 16 | (*a*) Give unadjusted estimates and, if applicable, confounder-adjusted estimates and their precision (eg, 95% confidence interval). Make clear which confounders were adjusted for and why they were included  (*b*) Report category boundaries when continuous variables were categorized  (*c*) If relevant, consider translating estimates of relative risk into absolute risk for a meaningful time period | Results – Smoking prevalence and intensity; Results – Educational inequalities; Results – Occupational inequalities; Results – Income-related inequalities; Table 2; Figures 1-3 | All paragraphs |
| Other analyses | 17 | Report other analyses done—eg analyses of subgroups and interactions, and sensitivity analyses | Results – Robustness checks and exploratory analyses; Appendix 3 & 4 | All paragraphs |
| **Discussion** | | |  |  |
| Key results | 18 | Summarise key results with reference to study objectives | Discussion | Paragraph 1 |
| Limitations | 19 | Discuss limitations of the study, taking into account sources of potential bias or imprecision. Discuss both direction and magnitude of any potential bias | Discussion | Paragraph 5 |
| Interpretation | 20 | Give a cautious overall interpretation of results considering objectives, limitations, multiplicity of analyses, results from similar studies, and other relevant evidence | Discussion | Paragraph 2-5 |
| Generalisability | 21 | Discuss the generalisability (external validity) of the study results | Discussion | Paragraph 2-5 |
| **Other information** | | |  |  |
| Funding | 22 | Give the source of funding and the role of the funders for the present study and, if applicable, for the original study on which the present article is based | Funding | n/a |

### Appendix 2 – Supplementary Methods

In this section methodological procedures are described in additional detail to provide interested readers with the depth needed to understand all details of the performed analyses.

#### 1. Collection of smoking behavior in the German Socioeconomic Panel over time

***Smoking status***

In 1998 smoking status was elicited by means of the following question:
*Do you currently smoke, whether cigarettes, a pipe, or cigars?*

- No
- Yes, primarily
  - Cigarettes
  - Pipes
  - Cigars

In 2001 smoking status was elicited by means of the following question:
*Do you smoke? Meant are cigarettes, pipes or cigars.*

- Yes, I am a smoker
- Was a smoker in the past
- Never smoked

From 2002 onwards smoking status was elicited in a consistent way by means of the following question with a disclaimer that information on e-cigarettes is collected elsewhere:
*Do you currently smoke, whether cigarettes, a pipe, or cigars?*

- Can`t / don`t want to answer
- Yes
- No

***Smoking intensity***

In 1998 smoking intensity was elicited by means of the following question:
*How many do you smoke approximately per day? If the number varies, please indicate the daily average for the past week.*

- On average, I smoke _____ cigarettes, pipes, cigars per day.

In 2001 smoking intensity was elicited by means of the following question:
*How many do you smoke approximately per day?*

- About _____ cigarettes, pipes, cigars.

For 1998 and 2001 we use the indicated number of smoked cigarettes, pipes and cigars as measure for smoking intensity.

From 2002 onwards smoking status was elicited in a consistent way by means of the following question:
*How many cigarettes, pipes or cigars do you smoke per day?*

- **Cigarettes:_____**
- Pipes:____
- Cigars / Cigarillos:______

We use the indicated number of smoked cigarettes as our measure for smoking intensity.

#### *Details on the occupational group classification*

To illustrate how the ISEI quintiles map onto concrete occupations, we report quintile boundaries and examples for occupations in each quintile for 2024 in this appendix.

Quintile boundaries for 2024 are:

- Q1 - Lowest: ISEI 11.6 to 28.5
- Q2 - Low: ISEI 28.5 to 50.4
- Q3 - Medium: ISEI 50.4 to 58.8
- Q4 - High: ISEI 58.8 to 74.7
- Q5 - Highest: ISEI 74.7 to 89.0

Below we list five example occupations per quintile to give an intuition for the kinds of jobs that fall within each range. The examples are common and well-known occupations in Germany; ISEI-08 scores for the corresponding ISCO-08 four-digit codes were taken from the Ganzeboom (2010) overview table [1].

Q1 – Lowest (*unskilled*):

- Crop farm labourer
- Building construction labourer
- Messenger / parcel deliverer
- Taxi / car driver
- Heavy truck / lorry driver

Q2 – Low (*semi-skilled manual*):

- Hairdresser
- Metal processing plant operator
- Nursing associate professional
- Bank teller
- Secretary (general)

Q3 – Medium (*skilled manual/lower white collar*):

- Sports coach / instructor
- Social work associate professional
- Administrative secretary
- Commercial sales representative
- Computer network technician

Q4 – High (*associate professional*):

- Nursing professional
- Primary school teacher
- Secondary school teacher
- Software developer
- Mechanical engineer

Q5 – Highest (*professional/managerial*):

- Architect
- University teacher
- Lawyer
- Generalist medical practitioner
- Specialist medical doctor

2. **Considerations for inequality measures**

Summary measures of socioeconomic inequality in health differ along three main axes: whether they quantify absolute or relative inequality, whether they use only the extremes of the socioeconomic distribution or its full range, and whether they are based on observed group values or on a fitted model [2]. The simplest measures contrast the two most extreme groups, namely the prevalence difference (absolute) and the prevalence ratio (relative). These are easy to communicate but draw on only two groups, ignore the distribution between them, depend strongly on how the extreme groups are defined, and disregard the relative size of each group [2].

The Slope Index of Inequality (SII) and Relative Index of Inequality (RII) overcome part of these limitations by regressing the outcome on the fractional rank of each group in the socioeconomic distribution, thereby using the entire distribution and accounting for group sizes. However, they assume a linear association between rank and outcome and extrapolate to the hypothetical individuals at the very bottom (rank 0) and top (rank 1) of the distribution, that is, beyond the observed group means. The ACI and RCI are derived from the covariance between the outcome and the same fractional rank [3,4]. They likewise use the full distribution and weight every socioeconomic position, but they do not extrapolate beyond the observed range. The RCI is bounded between -1 and +1, whereas the ACI is expressed on the scale of the outcome.

The regression-based and concentration-based families are closely related. For a given fractional rank R, the ACI equals $2\times Var(R)\times SII$ [4]. With equal-sized quintiles Var(R) is approximately 0.08, so the ACI is roughly 16% of the corresponding SII; for unequally sized categories such as the CASMIN educational groups the exact scaling factor differs. The numerically smaller ACI therefore reflects this scaling and not a smaller degree of inequality, and the two measures rank years and subgroups consistently.

We selected the ACI and RCI for three reasons: they account for all positions in the socioeconomic distribution more explicitly than the SII; they do not extrapolate beyond the observed distribution as regression-based measures do; and, in its absolute form, the index is expressed on the outcome scale and is therefore not rescaled by the declining mean prevalence. Because overall smoking prevalence fell markedly over the study period, relative measures can shift purely as a function of a shrinking mean, so we report the ACI alongside the RCI to distinguish changes in absolute magnitude from changes in the relative gradient. These properties make the ACI and RCI particularly suitable for comparisons over time [3,4]. Their main drawback is that they are less directly interpretable than simple group differences or ratios.

A further consideration arises because smoking status is binary. Wagstaff showed that for a binary outcome the concentration index cannot attain -1 and +1 but is bounded by (μ − 1) and (1 − μ), where μ is the mean prevalence; the attainable range therefore narrows as prevalence approaches 0 or 1, which can complicate comparisons across populations or time points with very different means [5]. The Wagstaff normalization rescales the index by 1/(1 − μ) to restore the full range. Erreygers subsequently proposed a corrected index for bounded variables that additionally satisfies the mirror property, meaning that inequality in smoking and in non-smoking are equal in magnitude and opposite in sign, as well as level independence [6].

We report the standard, uncorrected ACI and RCI for consistency and comparability with the established health-inequality literature. Because smoking prevalence in our population remains well away from 0 and 1 throughout the study period, the attainable range of the index stays wide, and the mean-dependence highlighted by Wagstaff has limited practical effect on our over-time and between-sex comparisons; reporting both the absolute and relative forms further guards against misinterpretation. Smoking intensity is a count rather than a binary variable, so the binary bound does not apply to the corresponding indices.

#### 3. Details on bootstrapping procedure

We estimate the sampling uncertainty of the Absolute and Relative Concentration Index (ACI / RCI) using a nonparametric percentile bootstrap with *B* = 1,000 replications. The procedure is implemented as a joint bootstrap stratified by survey year.

For each survey year *t*, we draw *B* bootstrap samples of size *n_t_* by resampling individuals with replacement from the year-specific cross-section. From each resample *b*, we compute the ACI and RCI for all outcome-subgroup combinations (prevalence and intensity; overall, male, and female) using the same resampled observations. This joint resampling ensures that the bootstrap distribution preserves the correlation structure across outcomes and subgroups.

For categorical socioeconomic indicators, such as educational level, the group assignments are fixed across resamples. For continuous indicators like equivalized household income and occupational status according to the Socio-Economic Index of Occupational Status (ISEI), we discretize into weighted quintiles, with quantile cut points recomputed within each bootstrap resample to correctly propagate the uncertainty arising from the discretization step into the confidence intervals [7].

When evaluating a subpopulation (e.g., smoking intensity among current smokers for men), the bootstrap resample is drawn from the full population and the socioeconomic ranking is computed on the full resample before applying the subset filter. This preserves the population-level rank ordering while restricting the outcome computation to the relevant subpopulation.

Confidence intervals (CI) are constructed as percentile intervals from the bootstrap distribution:

$$CI_{1-\alpha}=\left[ \hat{\theta}_{(\alpha/2)}^{*}, \hat{\theta}_{(1-\alpha/2)}^{*} \right]$$

where $\hat{\theta}_{(q)}^{*}$ denotes the *q*-th quantile of the *B* bootstrap estimates. To compute 95%-CIs, we set $\alpha$ to 5%. All analyses use cross-sectional survey weights throughout

### Appendix 3: Supplementary Tables

This section contains additional tables that support the main analysis and report results not directly available in the main manuscript.

#### Table A1 – Percentage of missing values across analysis variables and survey waves

| **Variable** | **Overall (all waves)** | **1998** | **2001** | **2002** | **2004** | **2006** | **2008** | **2010** | **2012** | **2014** | **2016** | **2018** | **2020** | **2022** | **2024** |
| --- | --- | --- | --- | --- | --- | --- | --- | --- | --- | --- | --- | --- | --- | --- | --- |
| Age | 0.00% | 0.00% | 0.00% | 0.00% | 0.00% | 0.00% | 0.00% | 0.00% | 0.00% | 0.00% | 0.00% | 0.00% | 0.00% | 0.00% | 0.00% |
| Sex | 0.02% | 0.00% | 0.00% | 0.00% | 0.00% | 0.00% | 0.00% | 0.00% | 0.00% | 0.00% | 0.00% | 0.00% | 0.02% | 0.09% | 0.07% |
| Migration Background | 0.00% | 0.00% | 0.00% | 0.00% | 0.00% | 0.00% | 0.00% | 0.00% | 0.00% | 0.00% | 0.00% | 0.00% | 0.00% | 0.00% | 0.00% |
| Relationship Status | 0.13% | 0.00% | 0.00% | 0.00% | 0.00% | 0.01% | 0.00% | 0.00% | 0.08% | 0.02% | 0.01% | 0.01% | 0.10% | 0.86% | 0.49% |
| Educational Level | 5.27% | 15.03% | 0.23% | 0.37% | 0.46% | 0.94% | 0.97% | 1.33% | 1.66% | 1.90% | 1.75% | 2.04% | 3.72% | 14.15% | 24.13% |
| Income (ENHI) | 0.08% | 0.00% | 0.00% | 0.00% | 0.00% | 0.00% | 0.00% | 0.00% | 0.00% | 0.00% | 0.00% | 0.01% | 0.86% | 0.00% | 0.00% |
| Occupational Status (ISEI) | 16.14% | 11.10% | 16.75% | 12.91% | 9.67% | 11.88% | 8.24% | 6.45% | 11.78% | 22.08% | 14.55% | 15.09% | 19.97% | 37.82% | 14.13% |
| Smoking Status | 0.21% | 0.26% | 0.34% | 0.04% | 0.24% | 0.09% | 0.11% | 0.11% | 0.32% | 0.15% | 0.13% | 0.11% | 0.17% | 0.35% | 0.42% |
| Smoking Intensity | 0.17% | 0.22% | 0.23% | 0.00% | 0.05% | 0.00% | 0.00% | 0.00% | 0.00% | 0.00% | 0.00% | 0.00% | 0.00% | 0.78% | 0.82% |
| Note: Values represent the percentage of missing observations per variable and survey wave. ISEI: Socio-Economic Index of Occupational Status | | | | | | | | | | | | | | | |

#### Table A2 – Age-standardized absolute and relative socioeconomic inequalities in smoking prevalence for 1998 and 2024

|  | **1998** | | **2024** | |
| --- | --- | --- | --- | --- |
| **Variable** | **Age-standardized** | **Crude** | **Age-standardized** | **Crude** |
| **Education** |  |  |  |  |
| No Graduation or Basic | 41.3% [39.5%–43.1%] | 39.2% [37.3%–41.1%] | 34.2% [30.5%–37.8%] | 33.5% [30.0%–37.0%] |
| Intermediate | 31.7% [29.0%–34.4%] | 36.9% [34.4%–39.3%] | 27.1% [24.4%–29.8%] | 27.3% [24.7%–29.9%] |
| Higher | 32.6% [26.1%–39.0%] | 36.7% [32.0%–41.4%] | 22.8% [19.8%–25.8%] | 22.7% [19.6%–25.7%] |
| Academic | 23.9% [21.1%–26.6%] | 24.6% [21.8%–27.4%] | 12.0% [10.4%–13.7%] | 11.3% [9.8%–12.8%] |
| **Occupational Status (ISEI)** |  |  |  |  |
| 1. Quintile – lowest | 41.7% [39.4%–44.1%] | 42.7% [40.3%–45.0%] | 34.7% [31.9%–37.5%] | 34.6% [31.9%–37.4%] |
| 2. Quintile | 41.0% [38.0%–44.1%] | 42.4% [39.5%–45.4%] | 28.4% [25.5%–31.3%] | 28.4% [25.5%–31.3%] |
| 3. Quintile | 34.4% [31.5%–37.2%] | 36.4% [33.5%–39.2%] | 20.0% [17.5%–22.5%] | 20.0% [17.5%–22.4%] |
| 4. Quintile | 29.7% [27.1%–32.3%] | 31.4% [28.6%–34.1%] | 16.6% [14.5%–18.6%] | 16.2% [14.1%–18.2%] |
| 5. Quintile – highest | 27.2% [24.4%–30.0%] | 28.2% [25.3%–31.0%] | 9.1% [7.4%–10.8%] | 9.0% [7.3%–10.6%] |
| **Equivalized Income** |  |  |  |  |
| 1. Quintile – lowest | 40.8% [38.2%–43.4%] | 42.5% [39.9%–45.1%] | 31.6% [29.0%–34.3%] | 30.9% [28.2%–33.5%] |
| 2. Quintile | 33.9% [31.4%–36.3%] | 35.2% [32.7%–37.6%] | 27.3% [24.8%–29.8%] | 27.2% [24.8%–29.7%] |
| 3. Quintile | 33.7% [31.2%–36.2%] | 34.5% [32.0%–37.0%] | 23.0% [20.5%–25.4%] | 22.9% [20.4%–25.4%] |
| 4. Quintile | 31.4% [28.9%–33.8%] | 32.1% [29.6%–34.6%] | 17.3% [15.2%–19.4%] | 17.5% [15.3%–19.8%] |
| 5. Quintile – highest | 31.0% [28.4%–33.7%] | 30.9% [28.3%–33.6%] | 12.0% [10.4%–13.6%] | 12.4% [10.7%–14.0%] |

**Notes:** The table presents age standardized and crude smoking prevalence for 1998 and 2024 stratified by education, occupational status and income and the corresponding 95%-Cis for the German population aged between 25 and 64. For the age standardization the German population from 2024 was used as standard population. CI: Confidence Interval. ISEI: Socio-Economic Index of Occupational Status

#### Table A3 – Absolute and relative socioeconomic inequalities in smoking prevalence and intensity for 1998 and 2024 for women

|  | **Smoking prevalence** | | | | |  | **Smoking intensity** | | | | |
| --- | --- | --- | --- | --- | --- | --- | --- | --- | --- | --- | --- |
|  | **1998** | | **2024** | | **Difference** |  | **1998** | | **2024** | | **Difference** |
|  | **Estimate** | **95% - CI** | **Estimate** | **95% - CI** | **2024 - 1998** |  | **Estimate** | **95% - CI** | **Estimate** | **95% - CI** | **2024 - 1998** |
| **Educational Level** |  |  |  |  |  |  |  |  |  |  |  |
| No graduation or Basic | 30.67% | [28.16%, 33.18%] | 34.82% | [29.45%, 40.19%] | 4.15% |  | 15.98 | [15.25, 16.71] | 14.48 | [13.35, 15.61] | -1.50 |
| Intermediate | 32.23% | [28.97%, 35.50%] | 24.99% | [21.59%, 28.40%] | -7.24% |  | 14.45 | [13.35, 15.54] | 12.00 | [10.79, 13.22] | -2.45 |
| Higher | 33.77% | [27.04%, 40.50%] | 20.72% | [16.36%, 25.08%] | -13.05% |  | 13.55 | [11.36, 15.74] | 11.08 | [9.84, 12.32] | -2.47 |
| Academic | 20.71% | [17.04%, 24.38%] | 10.10% | [7.91%, 12.28%] | -10.61% |  | 13.86 | [12.05, 15.67] | 10.33 | [8.15, 12.51] | -3.53 |
| Absolute inequalities (ACI) | -0.01 | [-0.02, -0.00] | -0.05 | [-0.06, -0.04] | -0.04 |  | -0.50 | [-0.78, -0.21] | -0.85 | [-1.24, -0.41] | -0.35 |
| Relative inequalities (RCI) | -0.03 | [-0.06, -0.00] | -0.24 | [-0.29, -0.20] | -0.21 |  | -0.03 | [-0.05, -0.01] | -0.07 | [-0.10, -0.03] | -0.04 |
| **Occupational Status (ISEI)** |  |  |  |  |  |  |  |  |  |  |  |
| 1. Quintile - lowest | 32.14% | [28.67%, 35.61%] | 32.60% | [28.54%, 36.66%] | 0.46% |  | 15.59 | [14.64, 16.55] | 13.36 | [12.26, 14.47] | -2.23 |
| 2. Quintile - low | 29.38% | [24.67%, 34.10%] | 25.02% | [20.87%, 29.18%] | -4.36% |  | 15.48 | [13.72, 17.24] | 11.76 | [10.57, 12.95] | -3.72 |
| 3. Quintile - medium | 33.96% | [30.28%, 37.63%] | 18.33% | [15.25%, 21.42%] | -15.63% |  | 14.98 | [13.85, 16.10] | 11.46 | [10.12, 12.81] | -3.52 |
| 4. Quintile - high | 30.66% | [27.23%, 34.10%] | 16.23% | [13.09%, 19.37%] | -14.43% |  | 15.02 | [13.86, 16.18] | 10.19 | [8.29, 12.08] | -4.83 |
| 5. Quintile - highest | 26.87% | [22.38%, 31.37%] | 6.43% | [4.49%, 8.38%] | -20.44% |  | 14.08 | [12.17, 16.00] | 9.62 | [7.26, 11.98] | -4.46 |
| Absolute inequalities (ACI) | -0.01 | [-0.02, 0.00] | -0.05 | [-0.06, -0.04] | -0.04 |  | -0.23 | [-0.56, 0.09] | -0.66 | [-1.04, -0.31] | -0.43 |
| Relative inequalities (RCI) | -0.02 | [-0.05, 0.01] | -0.23 | [-0.27, -0.20] | -0.21 |  | -0.02 | [-0.04, 0.01] | -0.06 | [-0.09, -0.03] | -0.04 |
| **Income** |  |  |  |  |  |  |  |  |  |  |  |
| 1. Quintile - lowest | 34.18% | [30.75%, 37.60%] | 26.70% | [23.34%, 30.05%] | -7.48% |  | 15.70 | [14.59, 16.80] | 12.81 | [11.56, 14.05] | -2.89 |
| 2. Quintile - low | 28.78% | [25.48%, 32.08%] | 24.34% | [21.03%, 27.64%] | -4.44% |  | 14.93 | [13.87, 15.98] | 12.48 | [11.27, 13.69] | -2.45 |
| 3. Quintile - medium | 28.45% | [25.06%, 31.84%] | 21.34% | [17.79%, 24.89%] | -7.11% |  | 14.06 | [12.97, 15.15] | 12.08 | [10.89, 13.27] | -1.98 |
| 4. Quintile - high | 25.69% | [22.35%, 29.04%] | 17.34% | [13.86%, 20.83%] | -8.35% |  | 16.39 | [14.98, 17.80] | 10.84 | [9.31, 12.38] | -5.55 |
| 5. Quintile - highest | 28.69% | [24.97%, 32.40%] | 9.55% | [7.45%, 11.66%] | -19.14% |  | 14.29 | [12.95, 15.62] | 11.97 | [9.85, 14.08] | -2.32 |
| Absolute inequalities (ACI) | -0.01 | [-0.02, -0.00] | -0.03 | [-0.04, -0.02] | -0.02 |  | -0.15 | [-0.47, 0.14] | -0.33 | [-0.67, 0.02] | -0.18 |
| Relative inequalities (RCI) | -0.04 | [-0.07, -0.01] | -0.16 | [-0.19, -0.12] | -0.12 |  | -0.01 | [-0.03, 0.01] | -0.03 | [-0.05, 0.00] | -0.02 |

**Notes:** The table presents smoking prevalence and smoking intensity measured by the number of consumed tobacco product per day stratified by education, occupational status and income, the corresponding 95%-Cis, ACIs and RCIs for the years 1998 and 2024 for the female German population aged between 25 and 64. The “Difference 2024 - 1998” column reports the respective prevalence/intensity/inequality measure difference between 2024 and 1998. ACI: Absolute Concentration Index. CI: Confidence Interval. RCI: Relative Concentration Index. ISEI: Socio-Economic Index of Occupational Status

#### Table A4 – Absolute and relative socioeconomic inequalities in smoking prevalence and intensity for 1998 and 2024 for men

|  | **Smoking prevalence** | | | | |  | **Smoking intensity** | | | | |
| --- | --- | --- | --- | --- | --- | --- | --- | --- | --- | --- | --- |
|  | **1998** | | **2024** | | **Difference** |  | **1998** | | **2024** | | **Difference** |
|  | **Estimate** | **95% - CI** | **Estimate** | **95% - CI** | **2024 - 1998** |  | **Estimate** | **95% - CI** | **Estimate** | **95% - CI** | **2024 - 1998** |
| **Educational Level** |  |  |  |  |  |  |  |  |  |  |  |
| No graduation or Basic | 47.57% | [44.88%, 50.26%] | 32.47% | [27.93%, 37.01%] | -15.10% |  | 20.87 | [20.02, 21.71] | 15.63 | [14.20, 17.06] | -5.24 |
| Intermediate | 42.29% | [38.53%, 46.05%] | 29.89% | [25.84%, 33.94%] | -12.40% |  | 18.07 | [17.02, 19.11] | 16.05 | [14.60, 17.49] | -2.02 |
| Higher | 39.61% | [33.04%, 46.17%] | 24.64% | [20.43%, 28.85%] | -14.97% |  | 16.04 | [13.92, 18.16] | 13.27 | [11.60, 14.94] | -2.77 |
| Academic | 27.40% | [23.41%, 31.39%] | 12.49% | [10.42%, 14.56%] | -14.91% |  | 16.31 | [14.55, 18.07] | 11.79 | [10.05, 13.52] | -4.52 |
| Absolute inequalities (ACI) | -0.04 | [-0.05, -0.03] | -0.05 | [-0.06, -0.04] | -0.01 |  | -1.00 | [-1.31, -0.68] | -0.82 | [-1.23, -0.37] | 0.18 |
| Relative inequalities (RCI) | -0.09 | [-0.12, -0.07] | -0.20 | [-0.24, -0.16] | -0.11 |  | -0.05 | [-0.07, -0.04] | -0.06 | [-0.09, -0.03] | -0.01 |
| **Occupational Status (ISEI)** |  |  |  |  |  |  |  |  |  |  |  |
| 1. Quintile - lowest | 50.67% | [47.52%, 53.82%] | 36.43% | [32.60%, 40.25%] | -14.24% |  | 21.10 | [20.17, 22.04] | 17.00 | [15.86, 18.13] | -4.10 |
| 2. Quintile - low | 48.20% | [44.55%, 51.85%] | 31.47% | [27.43%, 35.50%] | -16.73% |  | 19.11 | [18.10, 20.11] | 14.43 | [13.20, 15.66] | -4.68 |
| 3. Quintile - medium | 39.40% | [34.94%, 43.86%] | 22.60% | [18.49%, 26.71%] | -16.80% |  | 19.50 | [17.80, 21.20] | 12.99 | [11.12, 14.86] | -6.51 |
| 4. Quintile - high | 32.35% | [27.94%, 36.76%] | 16.14% | [13.40%, 18.87%] | -16.21% |  | 17.59 | [15.89, 19.29] | 10.94 | [9.19, 12.69] | -6.65 |
| 5. Quintile - highest | 28.94% | [25.27%, 32.60%] | 11.05% | [8.54%, 13.56%] | -17.89% |  | 17.02 | [15.52, 18.51] | 11.02 | [9.45, 12.60] | -6.00 |
| Absolute inequalities (ACI) | -0.05 | [-0.06, -0.04] | -0.05 | [-0.06, -0.05] | 0.00 |  | -0.80 | [-1.10, -0.46] | -1.31 | [-1.65, -0.92] | -0.51 |
| Relative inequalities (RCI) | -0.12 | [-0.14, -0.10] | -0.23 | [-0.26, -0.19] | -0.11 |  | -0.04 | [-0.06, -0.02] | -0.09 | [-0.12, -0.06] | -0.05 |
| **Income** |  |  |  |  |  |  |  |  |  |  |  |
| 1. Quintile - lowest | 51.95% | [48.11%, 55.79%] | 35.30% | [31.26%, 39.34%] | -16.65% |  | 18.83 | [17.67, 19.99] | 15.61 | [14.12, 17.10] | -3.22 |
| 2. Quintile - low | 41.95% | [38.36%, 45.53%] | 30.47% | [26.72%, 34.23%] | -11.48% |  | 19.21 | [18.06, 20.37] | 14.10 | [12.81, 15.38] | -5.11 |
| 3. Quintile - medium | 40.30% | [36.58%, 44.02%] | 24.45% | [21.01%, 27.89%] | -15.85% |  | 18.77 | [17.67, 19.86] | 13.79 | [12.42, 15.17] | -4.98 |
| 4. Quintile - high | 38.03% | [34.40%, 41.66%] | 17.75% | [14.96%, 20.54%] | -20.28% |  | 19.48 | [18.16, 20.80] | 14.13 | [13.00, 15.26] | -5.35 |
| 5. Quintile - highest | 32.86% | [29.13%, 36.59%] | 14.56% | [12.16%, 16.96%] | -18.30% |  | 19.82 | [18.34, 21.31] | 13.06 | [11.10, 15.01] | -6.76 |
| Absolute inequalities (ACI) | -0.03 | [-0.04, -0.02] | -0.04 | [-0.05, -0.04] | -0.01 |  | 0.18 | [-0.18, 0.47] | -0.44 | [-0.84, -0.06] | -0.62 |
| Relative inequalities (RCI) | -0.08 | [-0.11, -0.06] | -0.18 | [-0.21, -0.15] | -0.10 |  | 0.01 | [-0.01, 0.02] | -0.03 | [-0.06, -0.00] | -0.04 |

**Notes:** The table presents smoking prevalence and smoking intensity measured by the number of consumed tobacco product per day stratified by education, occupational status and income, the corresponding 95%-Cis, ACIs and RCIs for the years 1998 and 2024 for the male German population aged between 25 and 64. The “Difference 2024 - 1998” column reports the respective prevalence/intensity/inequality measure difference between 2024 and 1998. ACI: Absolute Concentration Index. CI: Confidence Interval. RCI: Relative Concentration Index. ISEI: Socio-Economic Index of Occupational Status

### Appendix 4: Supplementary Figures

This section contains additional figures that support the main analysis and report results not directly available in the main manuscript.

#### Figure A1 – Flow diagram for preparation of the analysis sample

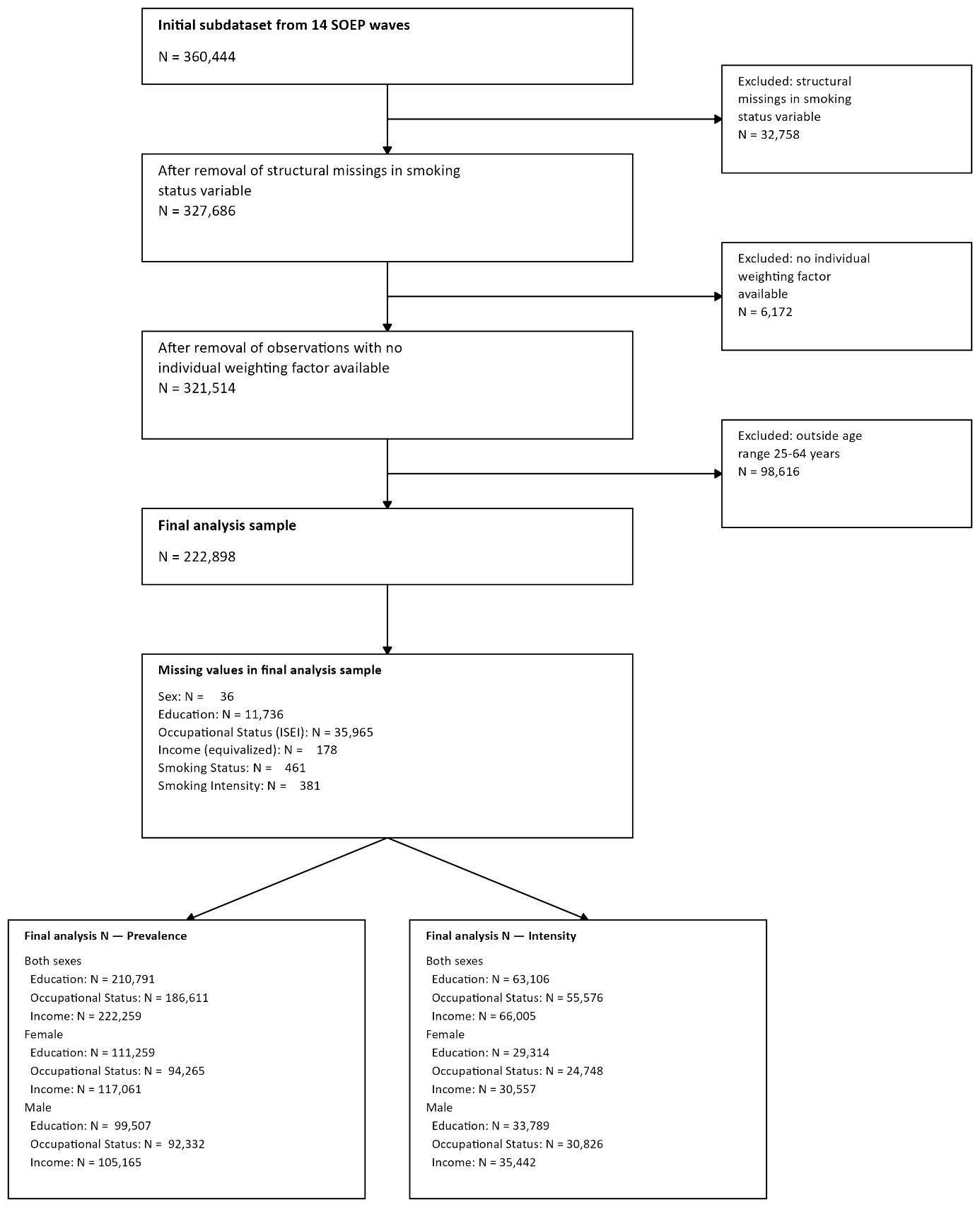

**Note:** Flowchart of the analysis sample construction from the initial 14-wave subdatasets (1998-2024). Boxes on the right summarize step-wise exclusions; boxes at the bottom report the number of missing values per variable in the final analysis sample and the resulting analysis Ns for the smoking prevalence and smoking intensity analyses, by sex and socio-economic status dimension.

#### Figure A2 – Smoking prevalence according to socioeconomic indicators from 1998 to 2024

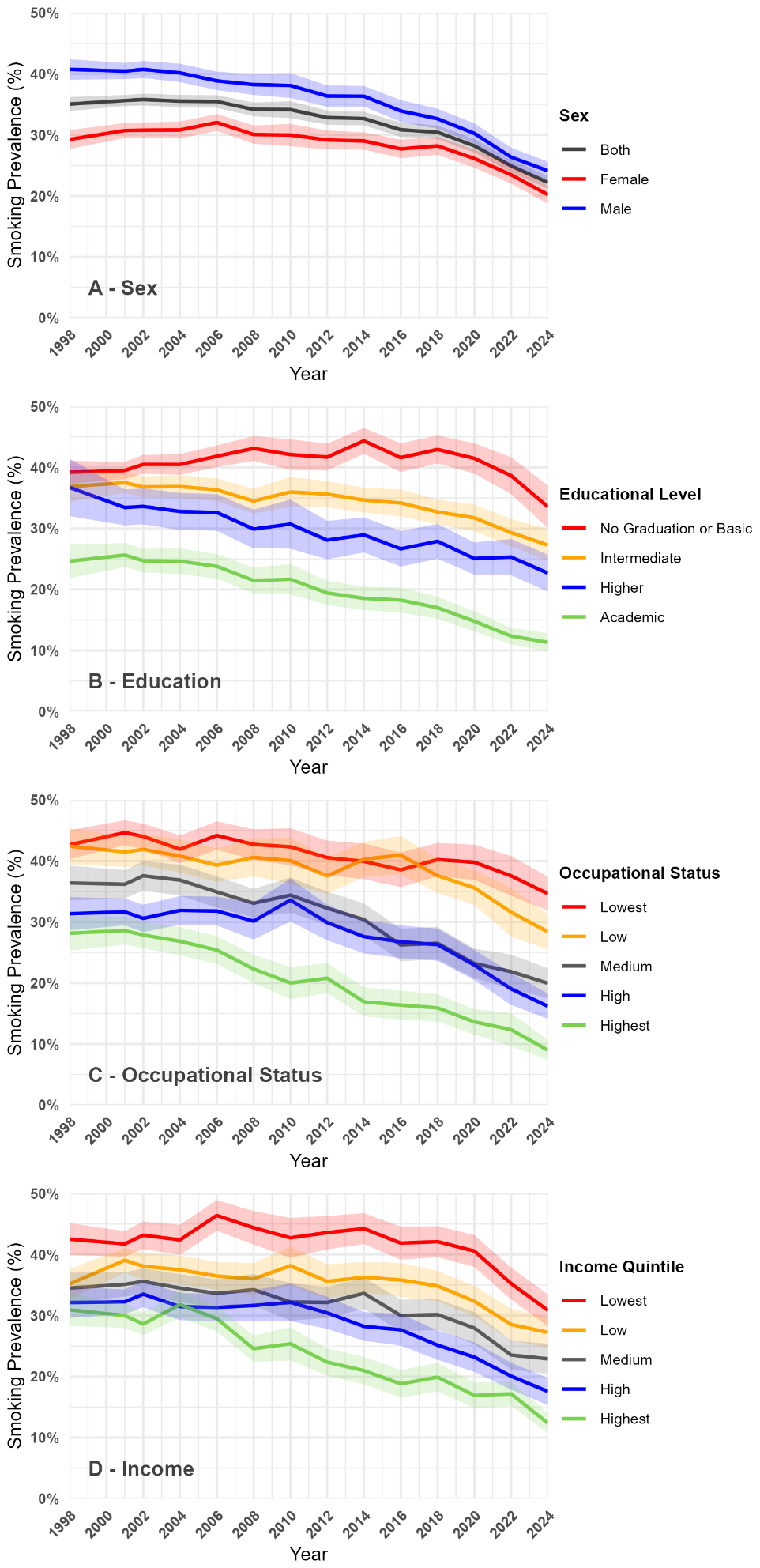

Note. Data: SOEP v41. Sample: adults aged 25-64. Lines show estimated smoking prevalence over time for the overall sample and separately for women and men. Shaded bands represent 95% confidence intervals. All estimates are weighted using the SOEP individual cross-sectional weighting factor.

#### Figure A3 – Smoking intensity according to socioeconomic indicators from 1998 to 2024

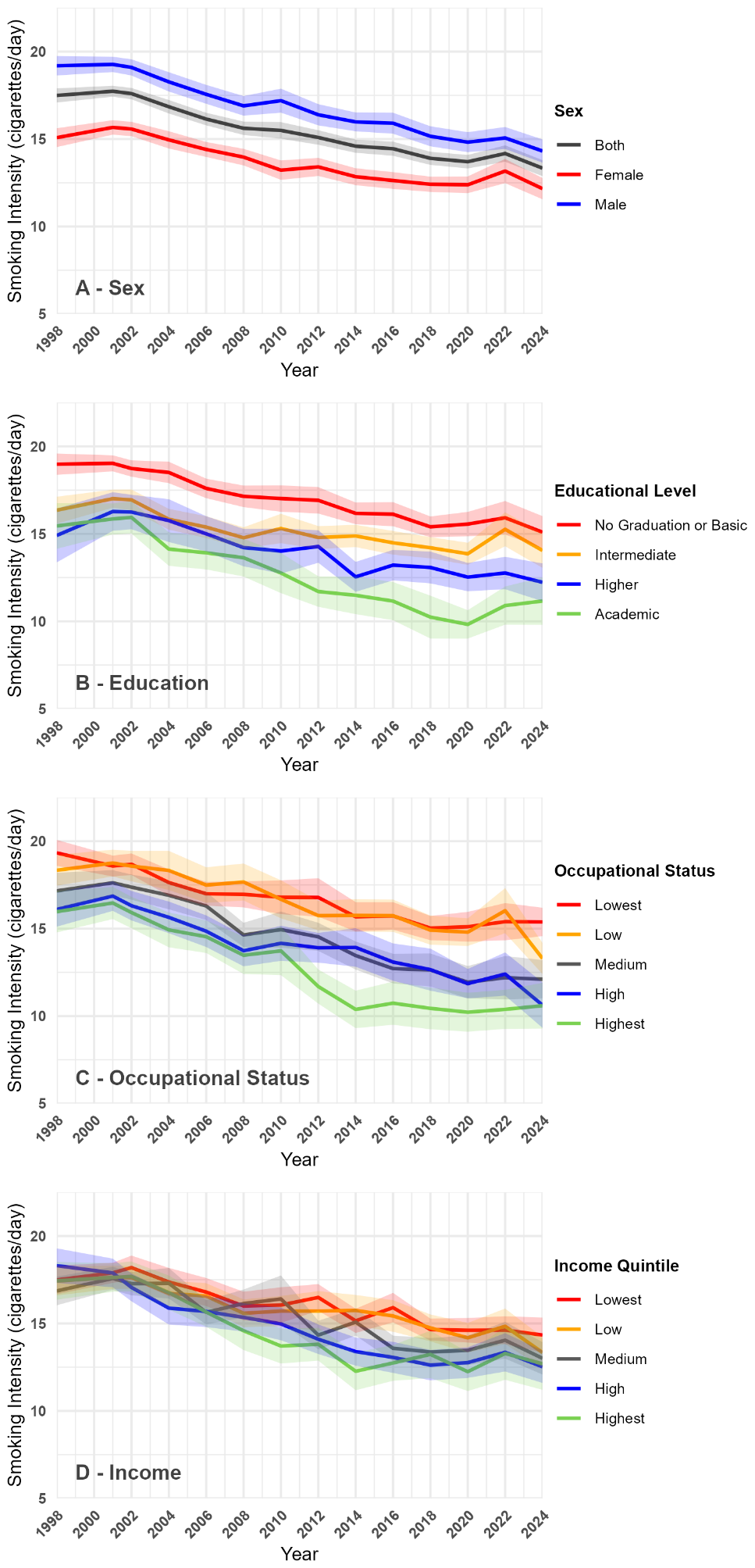

**Note:** Data: SOEP v41. Sample: current smokers aged 25-64. Lines show mean self-reported daily cigarette consumption (smoking intensity) over time for the overall sample and separately for women and men. Shaded bands represent 95% confidence intervals. All estimates are weighted using the SOEP individual cross-sectional weighting factor.

#### Figure A4 – Absolute and relative inequalities in smoking intensity according to socioeconomic indicators from 1998 to 2024 for men and women

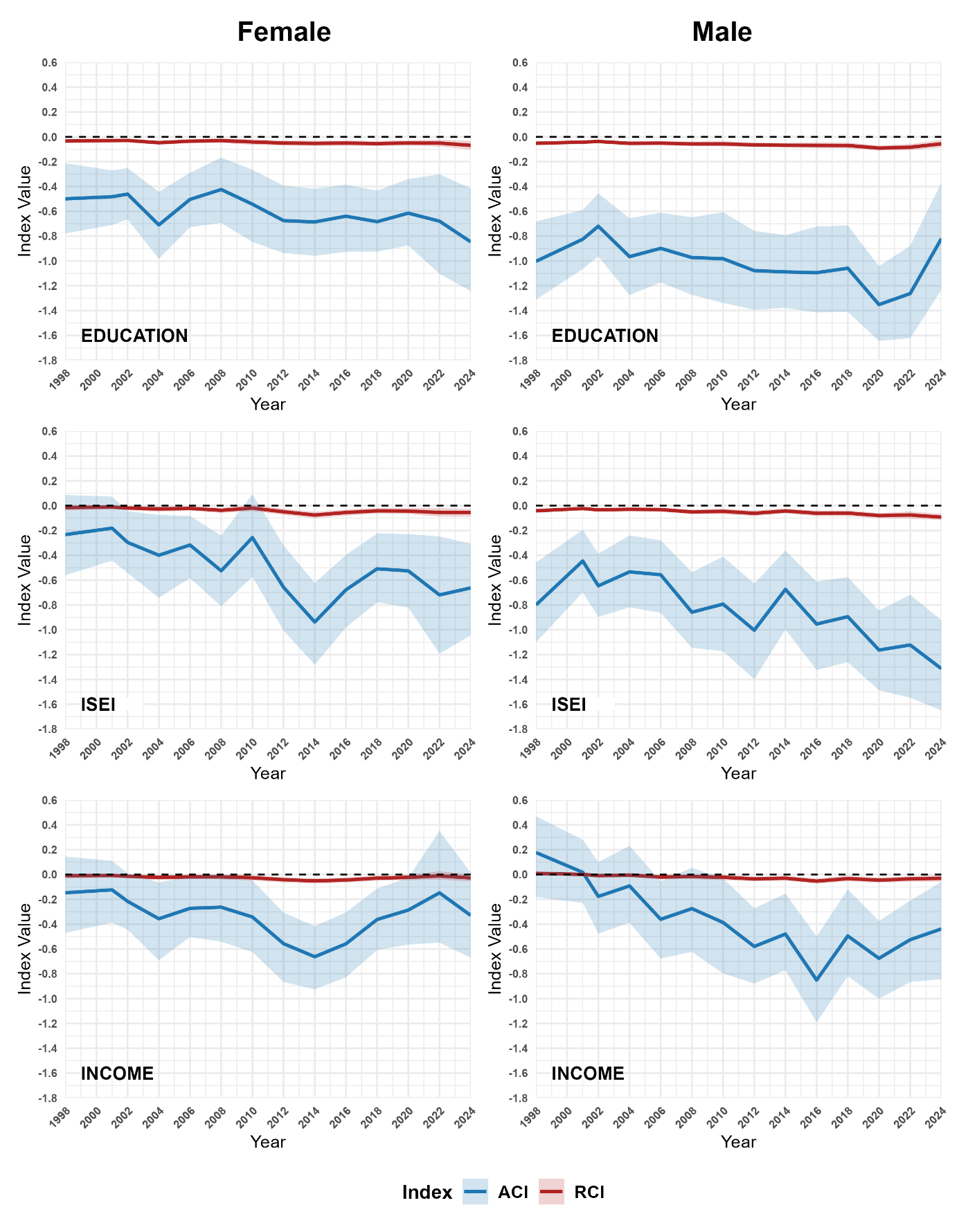

**Note:** Data: SOEP v41. Sample: current smokers aged 25-64. Analogous to Figure 3 for smoking intensity. Lines show the absolute (ACI) and relative (RCI) concentration index for smoking intensity by education, occupational status and income over time, separately for women and men. Negative values indicate higher smoking intensity among individuals with lower socio-economic status. Education levels follow the CASMIN classification (please see the methods section for details); occupational status (ISEI) and equivalized net household income (Income) are split into population-weighted quintiles for each survey year. Confidence intervals (shaded bands) are 95% percentile intervals from 1,000 bootstrap replications. All estimates are weighted using the SOEP individual cross-sectional weighting factor.

#### Figure A5 – Smoking prevalence according to education from 1998 to 2024 across of income quintiles

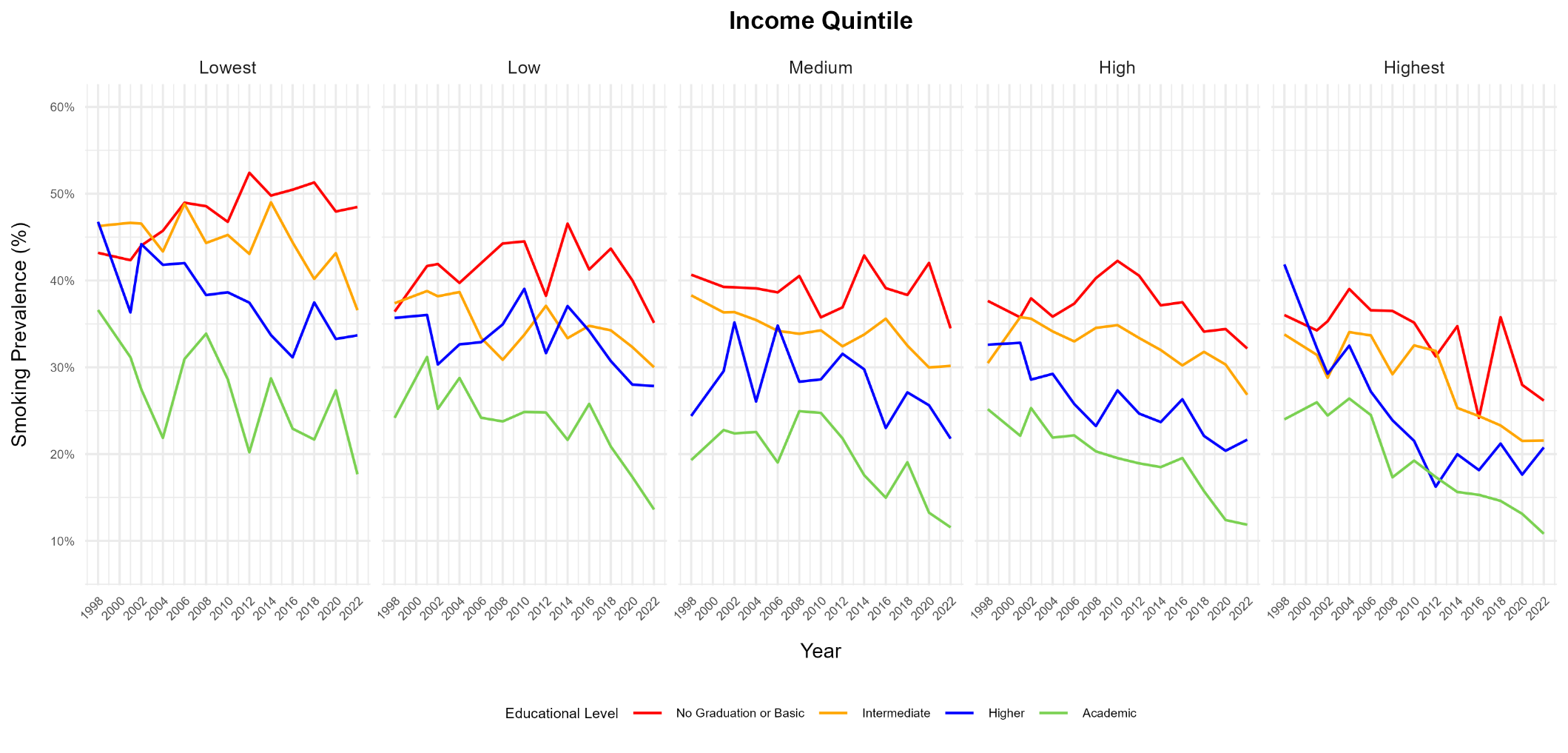

**Notes:** Data: SOEP v41. Sample: adults aged 25-64. Lines show estimated smoking prevalence by education level, stratified by income quintile, 1998-2024. Education levels follow the CASMIN classification (please see the methods section for details); equivalized net household income (Income) is split into population-weighted quintiles for each survey year. All estimates are weighted using the SOEP individual cross-sectional weighting factor.

#### Figure A6 – Smoking prevalence according to income quintiles from 1998 to 2024 across strata of education

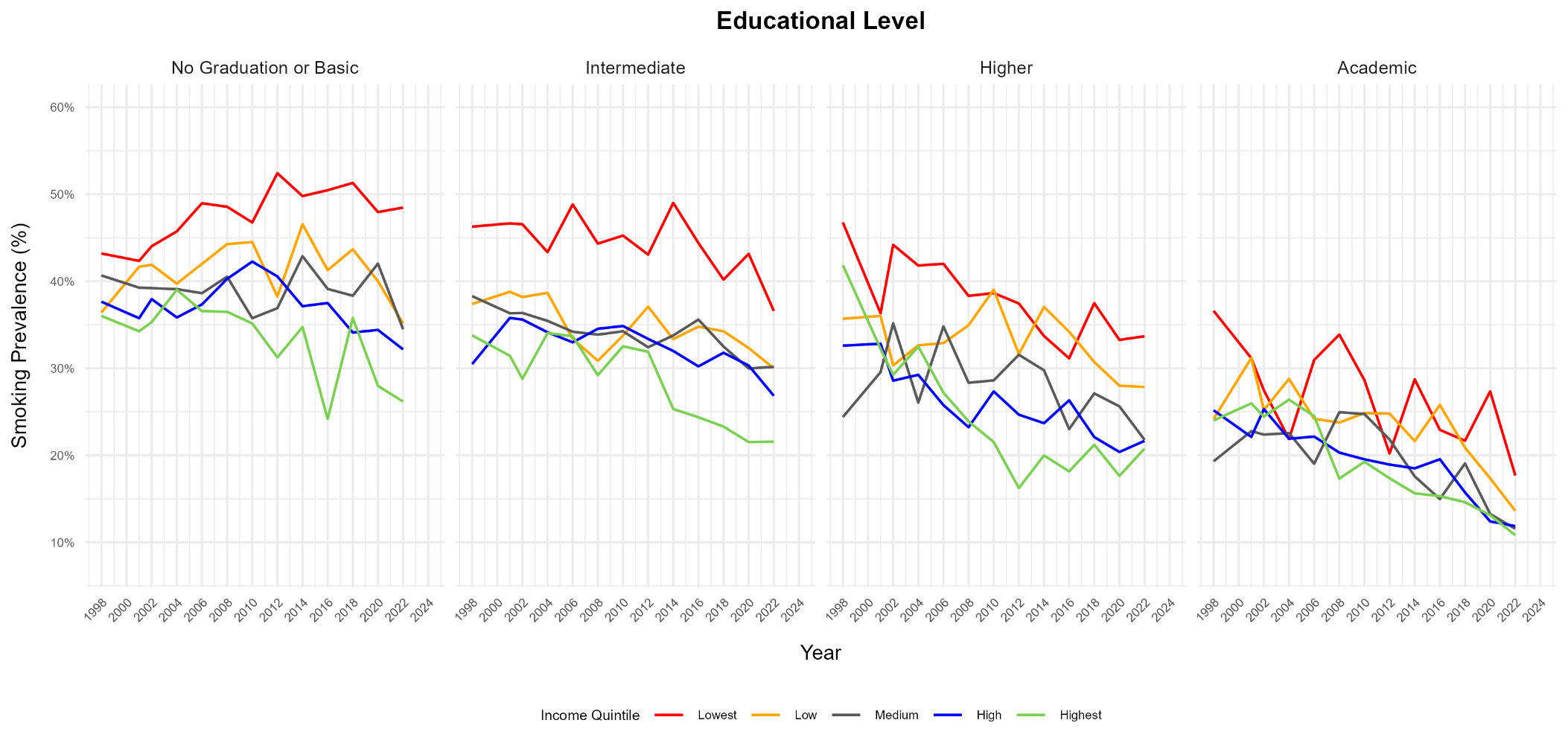

**Notes:** Data: SOEP v41. Sample: adults aged 25-64. Lines show estimated smoking prevalence by income quintile, stratified by education level, 1998-2024. Education levels follow the CASMIN classification (please see the methods section for details); equivalized net household income (Income) is split into population-weighted quintiles for each survey year. All estimates are weighted using the SOEP individual cross-sectional weighting factor.

#### Figure A7 – Smoking prevalence according to ISEI Qunitiles from 1998 to 2024 across Education levels

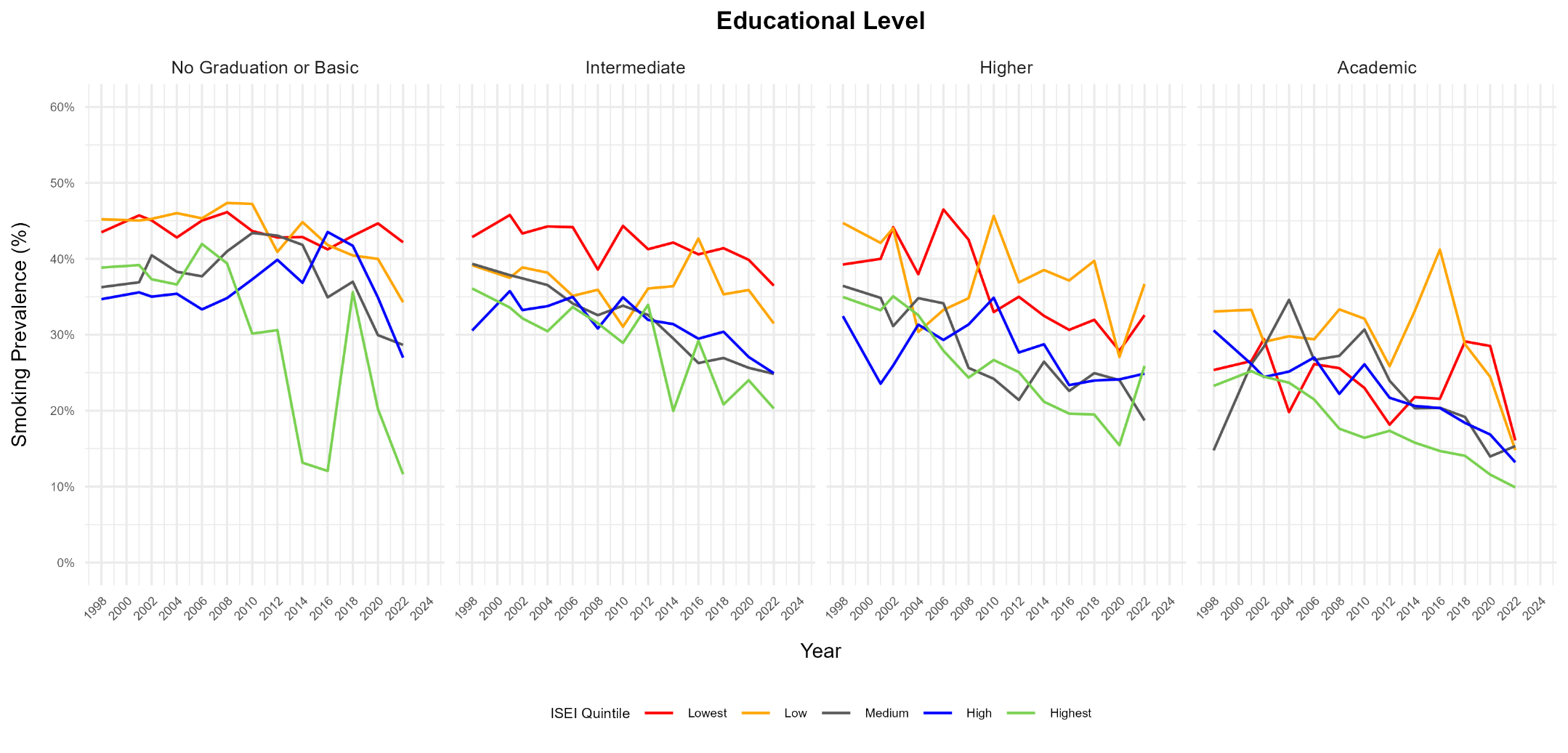

**Notes:** Data: SOEP v41. Sample: adults aged 25-64. Lines show estimated smoking prevalence by ISEI quintiles, stratified by education level, 1998-2024. Education levels follow the CASMIN classification (please see the methods section for details); occupational status (ISEI) is split into population-weighted quintiles for each survey year. All estimates are weighted using the SOEP individual cross-sectional weighting factor.

#### Figure A8 – Smoking prevalence according to education levels from 1998 to 2024 across ISEI quintiles

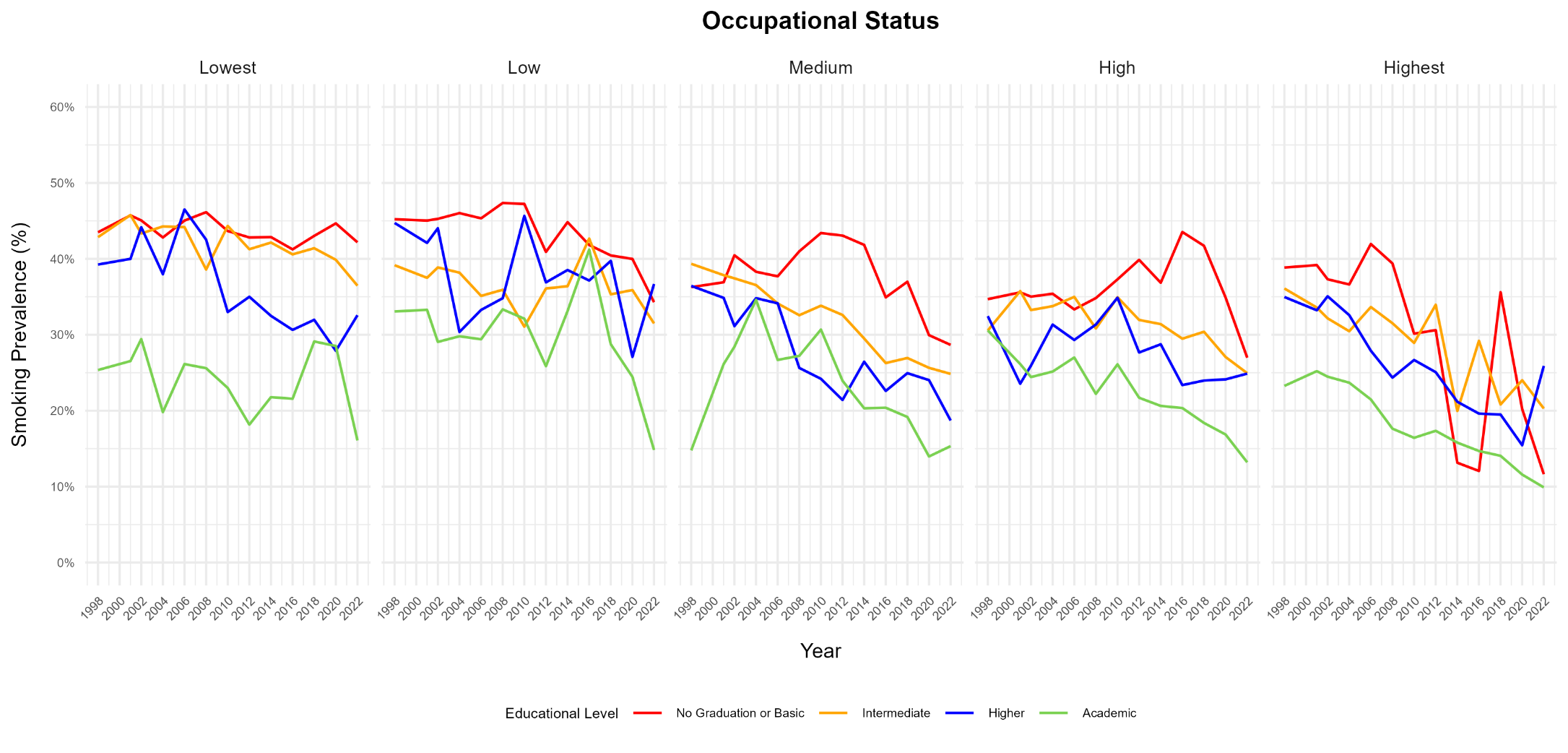

**Notes:** Data: SOEP v41. Sample: adults aged 25-64. Lines show estimated smoking prevalence by educational level, stratified by ISEI quintile, 1998-2024. Education levels follow the CASMIN classification (please see the methods section for details); occupational status (ISEI) is split into population-weighted quintiles for each survey year. All estimates are weighted using the SOEP individual cross-sectional weighting factor.

#### Figure A9 – Smoking prevalence according to income quintiles from 1998 to 2024 across ISEI quintiles

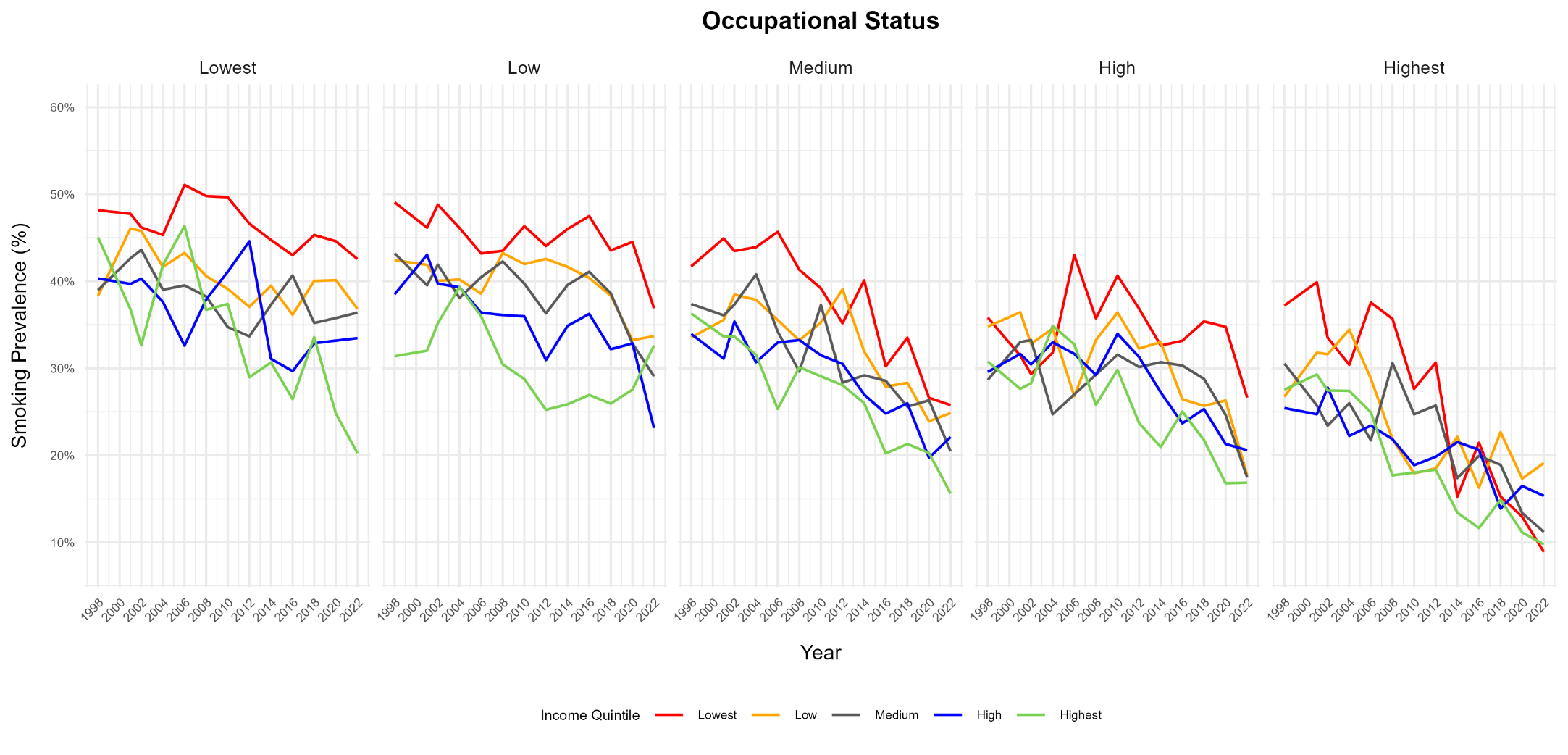

**Notes:** Data: SOEP v41. Sample: adults aged 25-64. Lines show estimated smoking prevalence by income quintile, stratified by ISEI quintile, 1998-2024. Occupational status (ISEI) and equivalized net household income (Income) are split into population-weighted quintiles for each survey year. All estimates are weighted using the SOEP individual cross-sectional weighting factor.

#### Figure A10 – Smoking prevalence according to ISEI quintiles from 1998 to 2024 across income quintiles

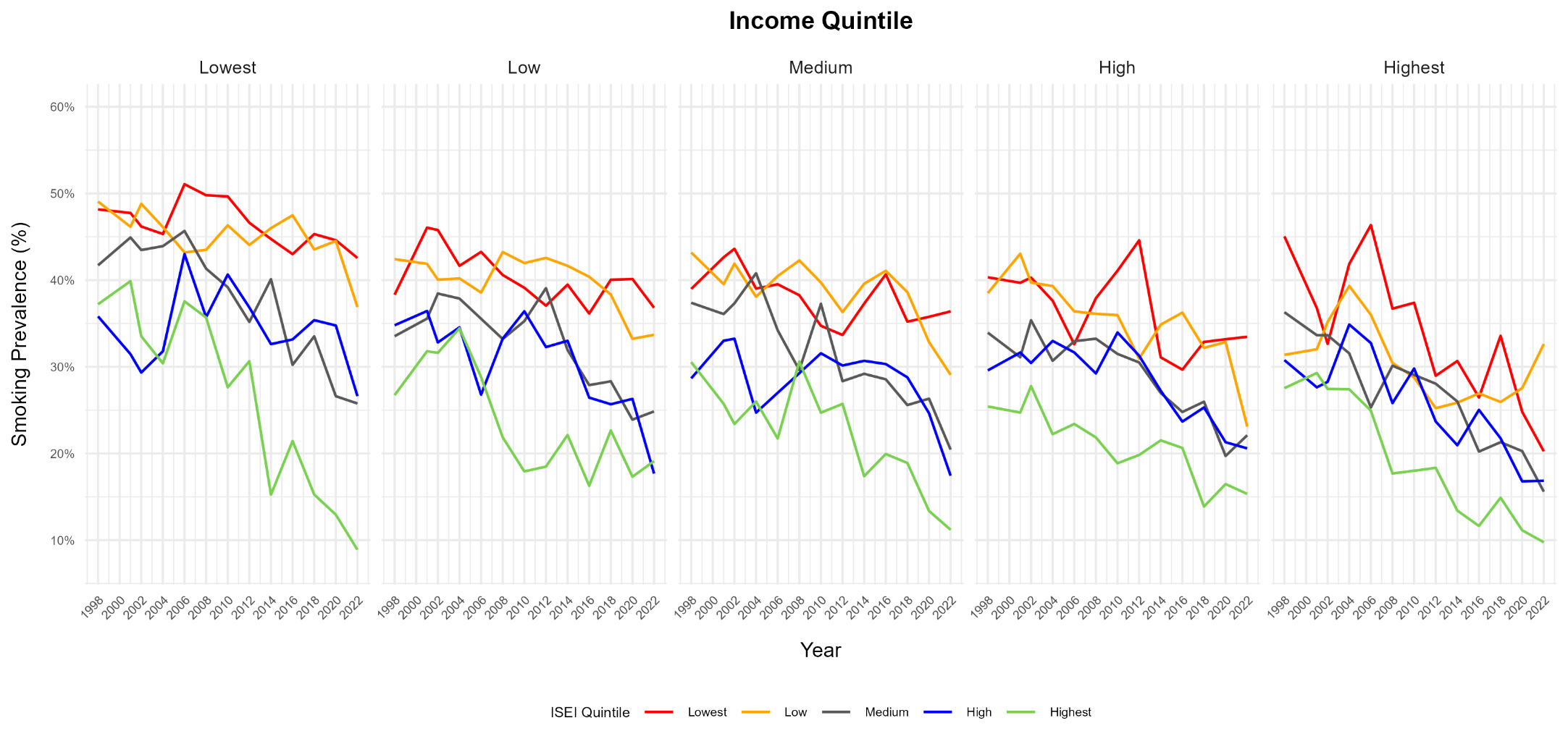
**Notes:** Data: SOEP v41. Sample: adults aged 25-64. Lines show estimated smoking prevalence by ISEI quintile, stratified by income quintile, 1998-2024. Occupational status (ISEI) and equivalized net household income (Income) are split into population-weighted quintiles for each survey year. All estimates are weighted using the SOEP individual cross-sectional weighting factor.

#### Figure A11 – Empirical density of equivalized household income in 2018, 2020, 2022, and 2024

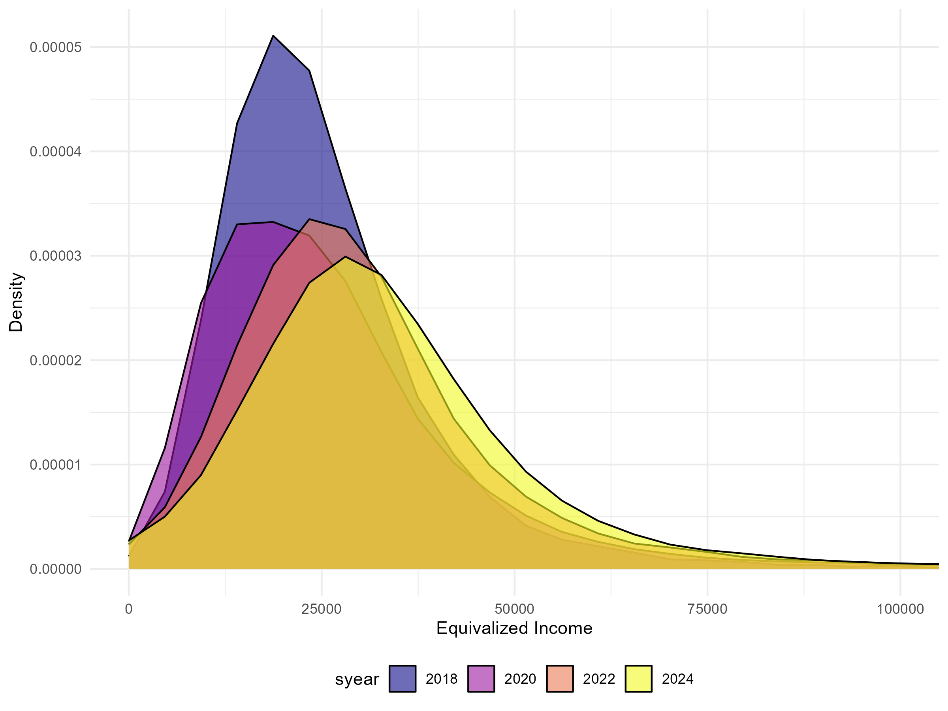

**Notes:** Data: SOEP v41. Sample: adults aged 25-64. Density of equivalized net household income in survey years 2018, 2020, 2022 and 2024.

#### Figure A12 – Histogram of equivalized household income in 2018, 2020, 2022, and 2024

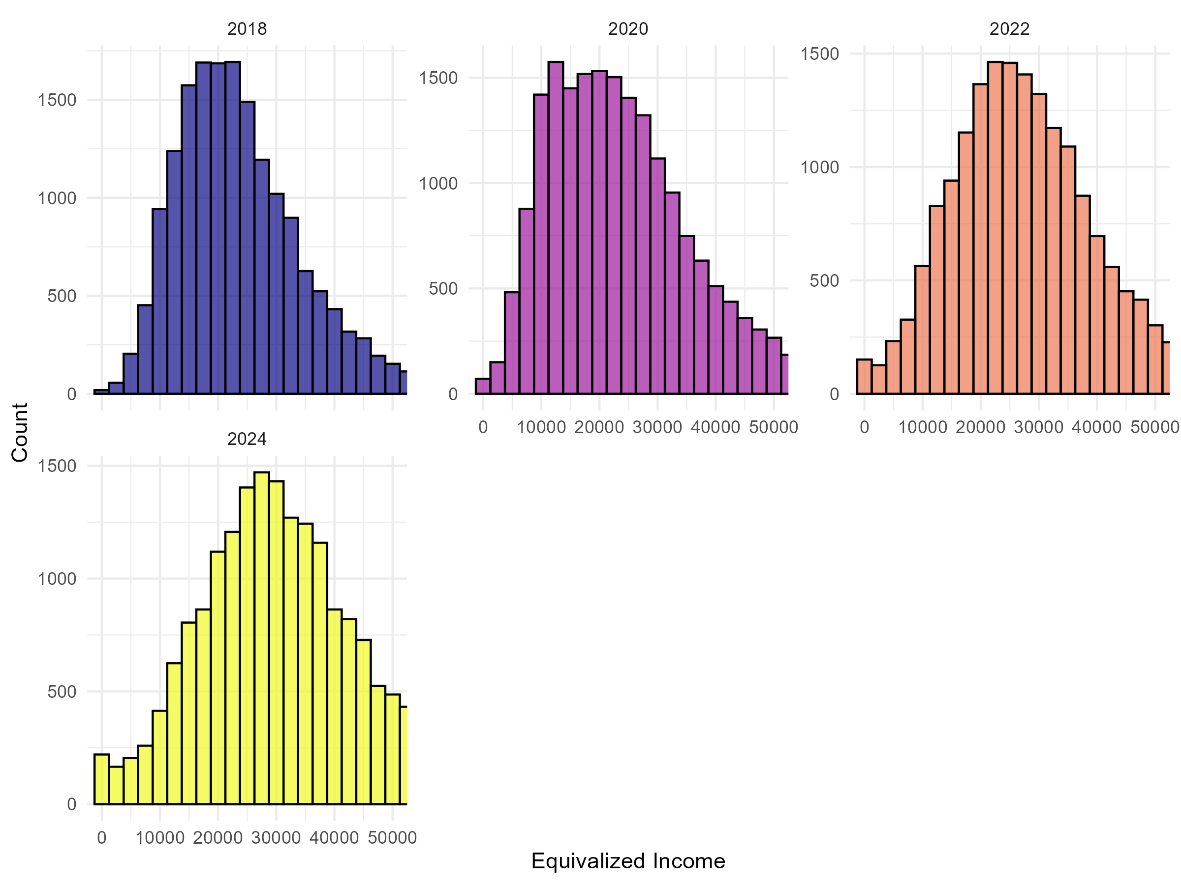

**Notes:** Data: SOEP v41. Sample: adults aged 25-64. Histogram of equivalized net household income (OECD-modified equivalence scale) in survey years 2018, 2020, 2022 and 2024.

#### Figure A13 – Distribution of educational level in the lowest income quintile in 2018, 2020, 2022, and 2024

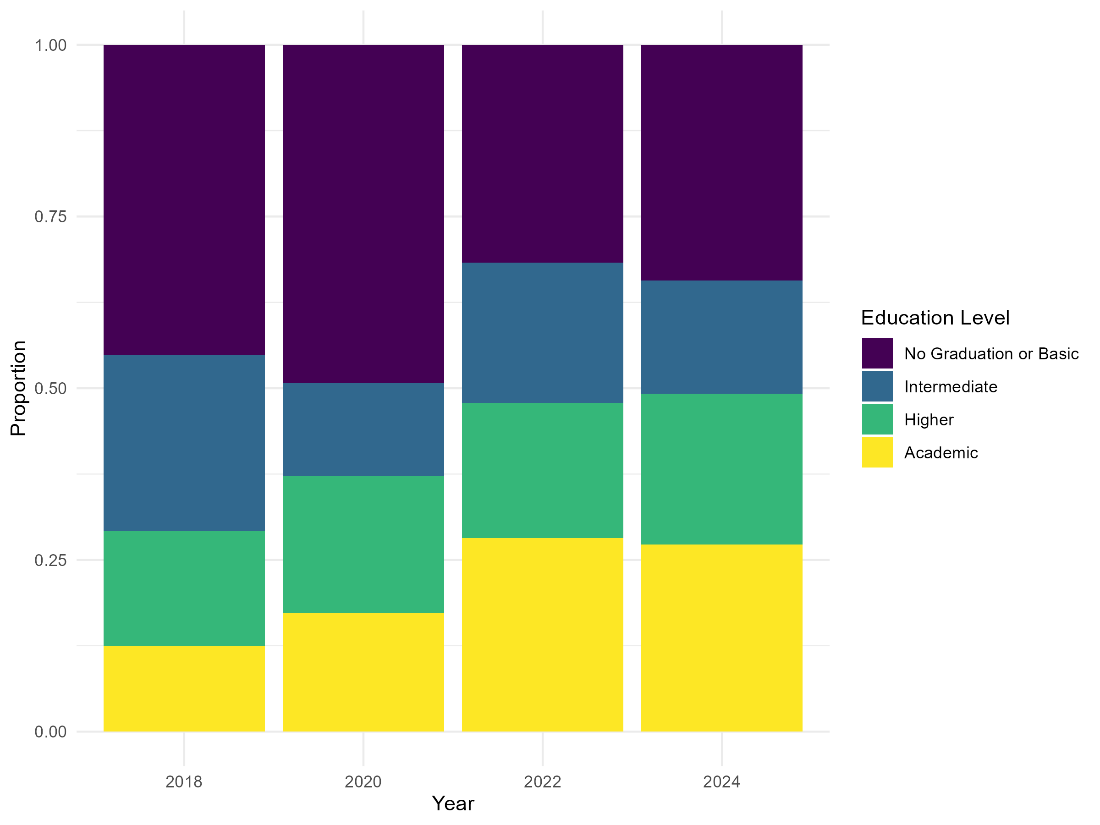

**Notes:** Data: SOEP v41. Sample: adults aged 25-64 in the lowest equivalized net household income quintile. Bars show the share of each education level within the lowest income quintile in survey years 2018, 2020, 2022 and 2024.

#### Figure A14 – Empirical density of age in the lowest income quintile in 2018, 2020, 2022, and 2024

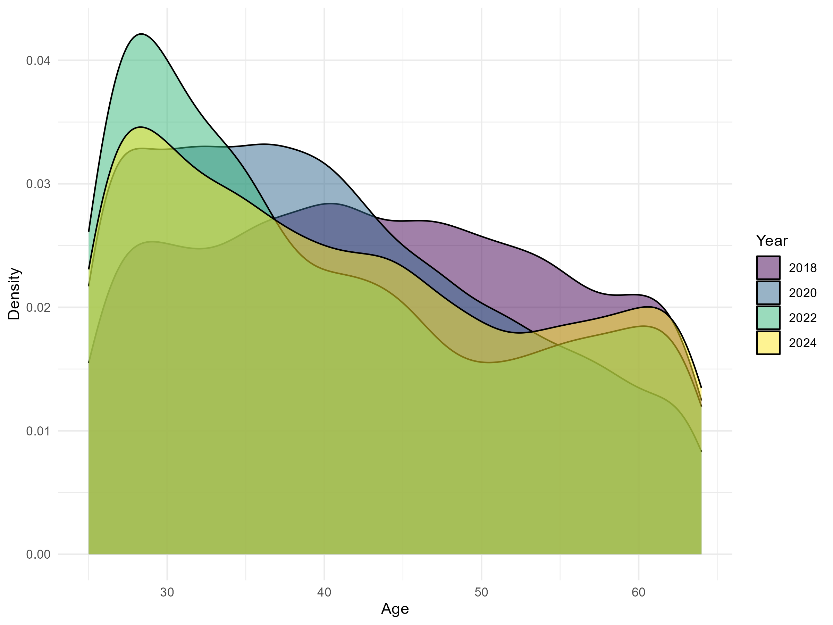

**Notes:** Data: SOEP v41. Sample: adults aged 25-64 in the lowest equivalized net household income quintile. The figure shows the age distribution within the lowest income quintile in survey years 2018, 2020, 2022 and 2024.
